# Success of prophylactic antiviral therapy for SARS-CoV-2: predicted critical efficacies and impact of different drug-specific mechanisms of action

**DOI:** 10.1101/2020.05.07.20092965

**Authors:** Peter Czuppon, Florence Débarre, Antonio Gonçalves, Olivier Tenaillon, Alan S. Perelson, Jérémie Guedj, François Blanquart

## Abstract

Repurposed drugs that are safe and immediately available constitute a first line of defense against new viral infections. Despite limited antiviral activity against SARS-CoV-2, several drugs are being tested as medication or as prophylaxis to prevent infection. Using a stochastic model of early phase infection, we find that there exists a critical efficacy that a treatment must reach in order to block viral establishment. For a single drug this efficacy is 87%, whereas for a combination of drugs this efficacy is reduced. Below the critical efficacy of any treatment, establishment of infection can sometimes be prevented, most effectively with drugs blocking viral entry into cells or enhancing viral clearance. Even when a viral infection cannot be prevented, antivirals delay the time to detectable viral loads. This delay flattens the within-host viral dynamic curve, possibly reducing transmission and symptom severity. Thus, antiviral prophylaxis, even with reduced efficacy, could be efficiently used to prevent or alleviate infection in people at high risk.

## 1 Introduction

The novel coronavirus SARS-CoV-2 rapidly spread around the globe in early 2020 (Li et al., 2020; Zhu et al., 2020; Lai et al., 2020; Chinazzi et al., 2020). As of November 13th, more than 50 million cases and 1.2 million associated deaths have been detected worldwide (Dong et al., 2020). SARS-CoV-2 causes substantial morbidity and mortality with about 4% of cases being hospitalized overall, but up to 47% in the oldest age group (Verity et al., 2020; Cereda et al., 2020; Salje et al., 2020), and a case fatality ratio of the order of 1% overall, which is again much higher in the elderly (Wu et al., 2020; Hauser et al., 2020; Verity et al., 2020). With a short epidemic doubling time of 2 to 7 days when uncontrolled (Li et al., 2020; Cereda et al., 2020; Muniz-Rodriguez et al., 2020), this epidemic can rapidly overburden healthcare systems (Ferguson et al., 2020). Many countries have imposed social distancing measures to reduce incidence. Lifting these measures while keeping the epidemic in check may require a combination of intensive testing, social isolation of positive cases, efficient contact tracing and isolation of contacts (Bi et al., 2020; Ferretti et al., 2020). Even if these measures are locally successful in keeping the disease at low prevalence, the presence of SARS-CoV-2 in many countries and substantial pre-symptomatic transmission (Tindale et al., 2020; Ferretti et al., 2020) suggest that the virus may continue to circulate for years to come.

Existing antiviral therapies can be repurposed to treat COVID-19 in infected individuals (Harrison, 2020; Li and Clercq, 2020; Gordon et al., 2020). Clinical trials to test several agents are underway, but existing antivirals have limited efficacy against SARS-CoV-2 and are most efficient in reducing viremia when taken early in infection (Gonçalves et al., 2020; Kim et al., 2020; Goyal et al., 2020). Prophylactic therapy using (repurposed) antivirals has been proposed (Jiang et al., 2020; Pagliano et al., 2020; Spinelli et al., 2020), is currently being tested (US National Library of Medicine, 2020 (accessed November 12, 2020) (e.g. study NCT04497987), and is successfully used in the prevention of HIV infection and malaria (Mermin et al., 2006; Baeten et al., 2012). Monoclonal antibodies, such as REGN-COV2 and Eli Lilly’s bamlanivimab, once authorized, could also be used for prophylaxis. These agents could be an essential tool to reduce the probability of SARS-CoV-2 infection in individuals at high risk, e.g. the elderly (especially those in nursing homes), individuals with co-morbidities, and health care workers, thus substantially reducing the burden on health care systems. Depending on the safety profile of the antiviral drug, it could be taken pre-exposure or just after contact with an infected individual (post-exposure). In this study, we integrate recent knowledge on SARS-CoV-2 host-pathogen interactions and the pharmacological properties of the antivirals currently tested in clinical trials to evaluate the efficacy of prophylactic antiviral therapy. We calculate the probability of establishment of a viral inoculum in an individual under prophylactic antiviral therapy.

## 2 Within-host model of viral dynamics

We consider a stochastic analog of a standard target-cell-limited model for viral kinetics. In this model, infectious virus particles, *V*_*I*_, infect target cells, *T*, i.e. cells susceptible to infection, in the upper respiratory tract at rate *β*. Initially, the resulting infected cells, *I*_1_, do not produce virus and are said to be in the eclipse phase of infection. After an average duration 1/*k*, these cells exit the eclipse phase and become productively infected cells, *I*_2_, which continuously produce virus at rate *p* per cell. A fraction *η* of these virions is infectious (*V*_*I*_) and can potentially infect new target cells s(*T*); the remainder of the produced virions, (1 −*η*), is non-infectious, denoted *V*_*NI*_. Non-infectious virions may be the result of deleterious mutations, or misassembly of the virus particle. Free virions (of both types) and infected cells are lost with rate *c* and *δ*, respectively. A potential early humoral immune response could contribute to the clearance parameter *c* or reduce the infection rate *β*. In other models, the innate immune response was assumed to increase the infected cell death rate *δ* (Goyal et al., 2020) or to reduce the number of available target cells by putting them into a refractory state (Pawelek et al., 2012; Gonçalves et al., 2020). It is currently not possible to decide on the best model structure to describe innate immunity given the limited available data during early infection. For large numbers of target cells, infected cells and virions, the following set of differential equations describes the dynamics:

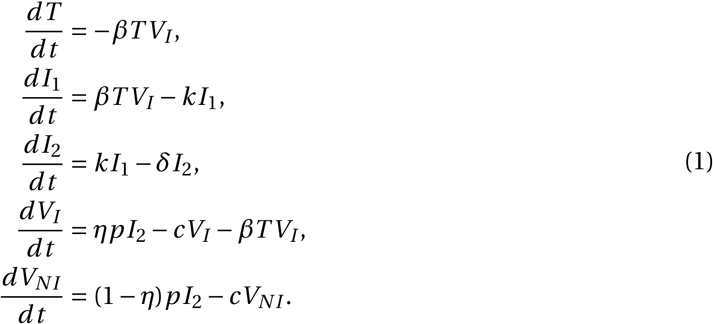

To generate parameter estimates for system (1), we followed the methodology of a previous study (Section S7 in the Supplementary Information (SI)) (Gonçalves et al., 2020). We show examples of our predictions in four out of 13 analyzed patients (Fig. 1a). An important quantity in determining the dynamics of this model is the within-host basic reproductive number *R*_0_. It reflects the mean number of secondary cell infections caused by a single infected cell at the beginning of the infection when target cells are not limiting. Using next-generation tools for invasion analysis (Hurford et al., 2010), the within-host basic reproductive number for model (1) is given by

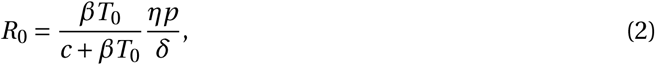

where *T*_0_ is the initial number of target cells. *R*_0_ is the product of two terms: *βT*_0_/(*c* + *βT*_0_), which corresponds to the probability that the virus infects a cell before it is cleared, and *ηp*/*δ*, which is the mean number of infectious virus particles produced by an infected cell during its lifespan of average duration 1/*δ*. The mean number of overall produced virions is called the “burst size” (*N = p*/*δ*). We study the within-host dynamics of SARS-CoV-2 in the early stage of an infection, when the number of infected cells is small and stochastic effects are important. To do so, we define a set of reactions corresponding to the differential equations in (1) (Pearson et al., 2011; Conway et al., 2013):

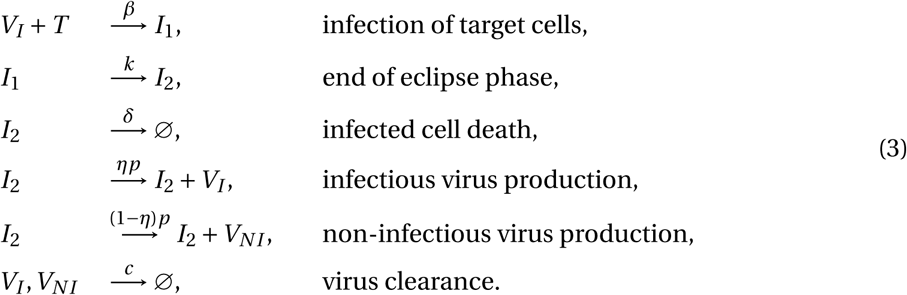

Because we are interested in early events, we subsequently assume in the analysis that the number of target cells remains equal to *T*_0_ (see Section S1 in the SI). This is a reasonable assumption as long as the number of virions is much smaller than the number of target cells (*V*_*I*_ (*t*) ≪ *T* (*t*)).

**Figure 1:**
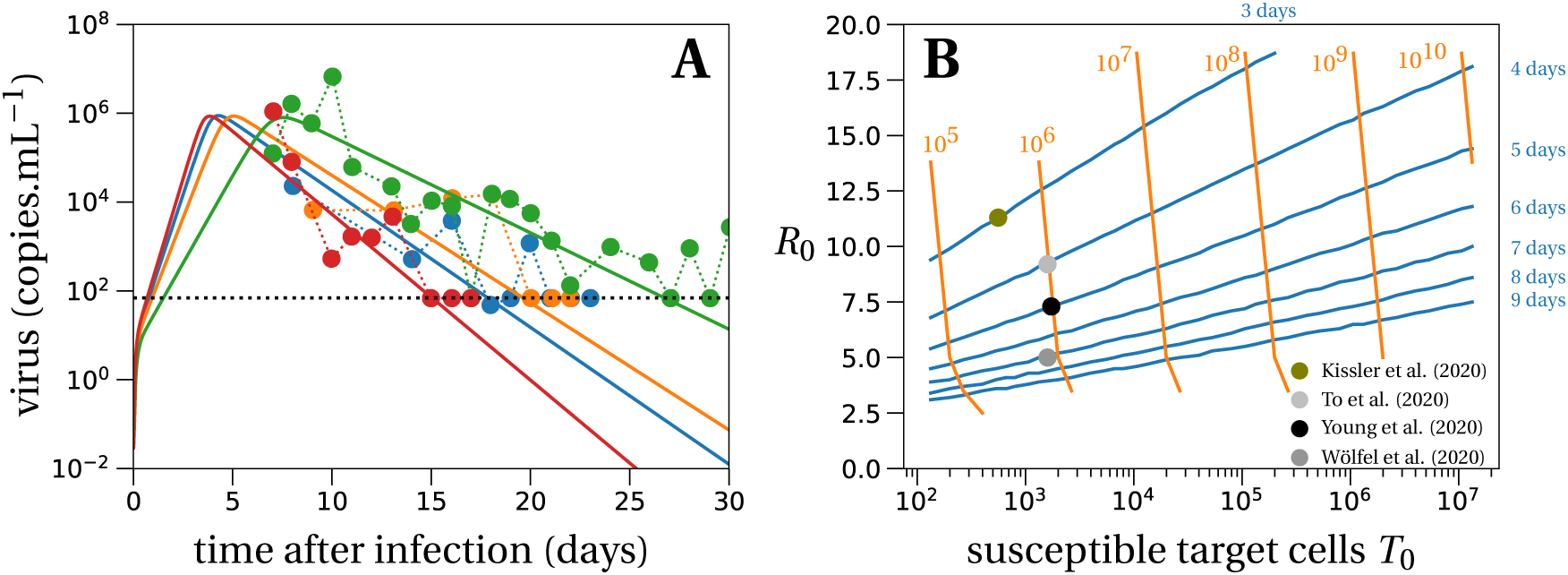
Deterministic within-host dynamics of SARS-CoV-2. (A) Model predictions using the target cell-limited model in four typical patients of Young et al. (2020). The estimated mean for within-host *R*_0_ of all patients from Young et al. (2020) is 7.69. Parameter values are given in Table S2 in the Supplementary Information. The dotted line depicts the detection threshold. (B) We plot the contour lines of the viral peak time (blue lines) and the number of virus particles at the viral peak per mL (orange lines) as a function of *R*_0_ and the number of susceptible target cells *T*_0_. The lines are obtained by evaluating the set of differential equations in Eq. (1) with different values of *T*_0_ (x-axis) and *R*_0_ (y-axis). The initial amount of virus particles per mL, *V*_*I*_ (0) = 1/30, corresponds to 1 infectious virus particle in absolute numbers in the total upper respiratory tract, which we assume has a volume of 30 mL. The contour lines for viral loads (orange) stop if the viral peak is reached later than 20 days after infection, which can happen for low values of within-host *R*_0_. The parameters of the model are set to: *k =* 5 day^−1^, *c =* 10 day^−1^, *δ =* 0.595 day^−1^, *p =* 11, 200 day^−1^, *η =* 0.001 and *β = cδR*_0_/(*T*_0_(*ηp* − *δR*_0_)) day^−1^. Dots depict averages of some data sets from Table 1.

### 2.1 Parameterization of the model

The exact values of the intrahost basic reproductive number *R*_0_ and the burst size *N* are critical to our predictions. Based on data from 13 patients (Young et al., 2020) with an observed peak viral load of order 10^6^ virions per mL, we estimate the intrahost basic reproductive number to be *R*_0_ = 7.69 (Fig. 1A), cf. Section S7 in the SI for more details and Gonçalves et al. (2020) for a sensitivity analysis of the same model without distinction of infectious and non-infectious virus. This sensitivity analysis revealed that intrahost *R*_0_ most likely lies somewhere between 2 and 18, in line with other estimates of *R*_0_ for SARS-CoV-2 in the upper respiratory tract (Ke et al., 2020). To further explore the range of *R*_0_ values compatible with other available data sets, we systematically solved the system of equations (1) and examined the peak viral load and the time when the peak is reached, as a function of the number of susceptible target cells *T*_0_ and *R*_0_, with all other parameters held constant at values given in Fig. 1B. For peak viral loads between 10^5^ and 10^8^ copies per mL and peak timing between 3 and 9 days, encompassing the range of average outcomes observed in multiple studies (Table 1), *R*_0_ may vary between 3 and 13 (Fig. 1B). We note that there is substantial inter-individual variability in viral loads, and some patients present an observed peak viral load at 10^9^ copies/mL or higher (Jones et al., 2020; Kissler et al., 2020), compatible with a *R*_0_ of 15 or more. The mean observed peak viral load across the studies surveyed was 10^6^ copies/mL (Table 1).

**Table 1:** Literature review of SARS-CoV-2 viral load trajectories within hosts.

| Country / Setting | # ind. | Mean observed peak viral load<br>[copies.mL<sup>-1</sup>] | Mean time of observed viral peak [days after infection] | Reference |
| --- | --- | --- | --- | --- |
| Singapore / hospital / nasopharyngeal swabs | 13 | 10<sup>6</sup><br>(max. 3 × 10<sup>8</sup>) | 5-10 (a few days after symptoms) | Young et al. (2020) |
| Germany / hospital / nasopharyngeal swabs | 9 | 7 × 10<sup>5</sup><br>(max. 2 × 10<sup>9</sup>) | ≤ 7 (already declining at admission) | Wölfel et al. (2020) |
| mainland China / throat swabs | 67 | 10<sup>5</sup><br>(max. 3 × 10<sup>7</sup>) | ≤ 5 (no increase after symptom onset) | Pan et al. (2020) |
| mainland China / throat swabs | 94 | 10<sup>5</sup><br>(max. 7 × 10<sup>8</sup>) | 5 | He et al. (2020) |
| Hong Kong / hospital / throat swabs | 23 | 10<sup>6</sup><br>(max. 3 × 10<sup>7</sup>) | 4 | To et al. (2020) |
| France / hospital / nasopharyngeal swabs | 25 | 6 × 10<sup>8</sup><br>(max. 2 × 10<sup>11</sup>) | 9<br>(inferred in prospective study) | Tubiana et al. (2020) |
| USA / NBA players and staff / nasopharyngeal and throat swabs | 68 | 4 × 10<sup>5</sup><br>(max. 10<sup>7</sup>) | 3 | Kissler et al. (2020)* |

Alongside the mean observed peak viral loads, we also state the maximal peak viral loads from the cited studies (minimal values are not always provided in the references). These maximal values inform about the plausible upper bound for the within-host reproductive number $R_0$ . \*Cycle threshold (Ct) values are reported. Conversion to viral loads is according to personal communication with David Ho (Columbia University).

The burst size for SARS-CoV-2 is unknown. Estimates of the burst size for other coronaviruses range from 10 − 100 (Robb and Bond, 1979) to 600 − 700 (Bar-On et al., 2020; Hirano et al., 1976) infectious virions. We assume that the proportion of infectious virions produced by an infected cell is *η =* 10^−3^. This value is motivated by the fraction of infectious virus in an inoculum injected into rhesus macaques, *η =* 1.33 × 10^−3^ (Munster et al., 2020). The total viral burst size is then between 10, 000 and 100, 000 virions. Such large total burst size is suggested by electron microscopy showing the emergence of many virions from cells infected by SARS-CoV-1 (Stertz et al., 2007; Knoops et al., 2008) (see also National Institute of Allergy and Infectious Diseases (NIAID) (2020 (accessed November 5, 2020), a webpage dedicated to SARS-CoV-2: e.g. https://www.flickr.com/photos/niaid/49557785797/in/album-72157712914621487/). Given the uncertainty in this parameter, we ran simulations with a small (parameter set ‘LowN’) and a large burst size (parameter set ‘HighN’). The exact values of the LowN and HighN parameter sets are given in Table 2.

**Table 2:** Model parameters used in the stochastic simulations.

| Parameter set | $\eta p$ [day<sup>-1</sup>] | $T_0$ [cells] | $\eta N$ [virions] | $R_0$ [cells] |
| --- | --- | --- | --- | --- |
| LowN | 11.2 | $4 \times 10^4$ | 18.8 | 7.69 |
| HighN | 112 | $4 \times 10^3$ | 188 | 7.69 |

Parameters not shown in the table are not changed between the simulations and are set to: $k = 5 \text{ day}^{-1}$ , $\delta = 0.595 \text{ day}^{-1}$ , $c = 10 \text{ day}^{-1}$ , $\eta = 10^{-3}$ , $\beta = c\delta R_0 / (T_0(\eta p - \delta R_0)) \text{ day}^{-1}$ .

## 3 Results

### 3.1 Survival and establishment of the virus within the host

As shown previously (Pearson et al., 2011; Conway et al., 2013), with the model dynamics defined in (3) the probability that a viral inoculum of size *V*_0_ establishes an infection within the host is given by:

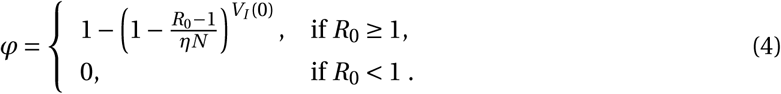

When *R*_0_ > 1, the establishment probability increases with the size of the inoculum *V*_*I*_ (0). Indeed, for infection to succeed, only a single infectious virus particle among *V*_*I*_ (0) needs to establish, so the more virus particles there are initially, the more likely it is that at least one establishes. Importantly, for a given *R*_0_, the virus establishes more easily when it has a low burst size *N*. Keeping the mean number of secondary cell infections *R*_0_ constant, a virus with a smaller burst size will have a larger infectivity *β* or smaller clearance *c*, which increases the first factor of *R*_0_ (Eq. (2)). For the same number of virions to be produced at lower burst sizes, more cells need to be involved in viral production than for large burst sizes. This mitigates two risks incurred by the virus: the risk that it does not find a cell to infect before it is cleared, and the risk that the infected cell dies early by chance. Since more cells are involved in viral production for lower burst sizes, these risks are shared over all these virus-producing cells. This reduces the stochastic variance in viral production, which in turn results in a higher establishment probability.

### 3.2 Prophylactic antiviral therapy blocks establishment of the virus

Next, we investigate the effect of prophylactic antiviral drug therapy on the establishment probability of the virus during the early phase of an infection. In particular, we examine drugs with four distinct modes of action.

i. Reducing the ability of the virus to infect cells *β*. This corresponds for instance to treatments that block viral entry, e.g. a neutralizing antibody (given as a drug) that binds to the spike glycoprotein (Chen et al., 2020).
ii. Increasing the clearance of the virus *c*. This mode of action models drugs such as monoclonal antibodies that may be non-neutralizing or neutralizing and bind to circulating virus particles and facilitate their clearance by phagocytic cells (Igarashi et al., 1999).
iii. Reducing viral production *p*. This mechanism corresponds for example to nucleoside analogues that prevent viral RNA replication (favipiravir, remdesivir), or to protease inhibitors (lopinavir/ ritonavir)(Li and Clercq, 2020).
iv. Increasing infected cell death *δ*. This would describe the effect of SARS-CoV-2 specific antibodies that bind to infected cells and induce antibody-dependent cellular cytoxicity or antibody-dependent cellular phagocytosis. It would also model immunomodulatory drugs that stimulate cell-mediated immune responses, or immunotoxins such as antibody toxin conjugates that can directly kill cells (Hoffmann et al., 2020).

We denote by *ε*_*β*_, *ε*_*c*_, *ε*_*p*_ and *ε*_*δ*_ the efficacies of the antiviral drugs in targeting the viral infectivity, viral clearance, viral production and infected cell death, respectively. Their values range from 0 (no efficacy) to 1 (full suppression). We neglect variations in drug concentrations over time within the host and, to be conservative, assume a constant drug efficacy corresponding to the drug efficacy at the drug’s minimal concentration between doses.

#### 3.2.1 Antiviral reducing viral infectivity

Antiviral drugs reducing viral infectivity *β* by the factor (1 −*ε*_*β*_) leave the burst size *N* unchanged, but reduce the basic reproductive number, *R*_0_, by a factor 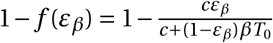. If (1 − *f*(*ε*_*β*_)) × *R*_0_ ≥ 1, the establishment probability changes to:

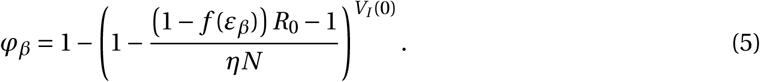

If 1 − *f* (*ε*_*β*_) × *R*_0_ is less than 1, the virus will almost surely go extinct and we have *φ*_*β*_ = 0.

With a plausible inoculum size of 10 infectious virions (Leung et al., 2020), we find that an efficacy (*ε*_*β*_) of 81% (LowN parameter set) is necessary to reduce the establishment probability of a viral infection by 50% compared to no treatment (see Fig. 2 panels A and C). Subsequently, when we mention the efficacy of an antiviral drug reducing viral infectivity, we always refer to *ε*_*β*_ and not *f* (*ε*_*β*_).

**Figure 2:**
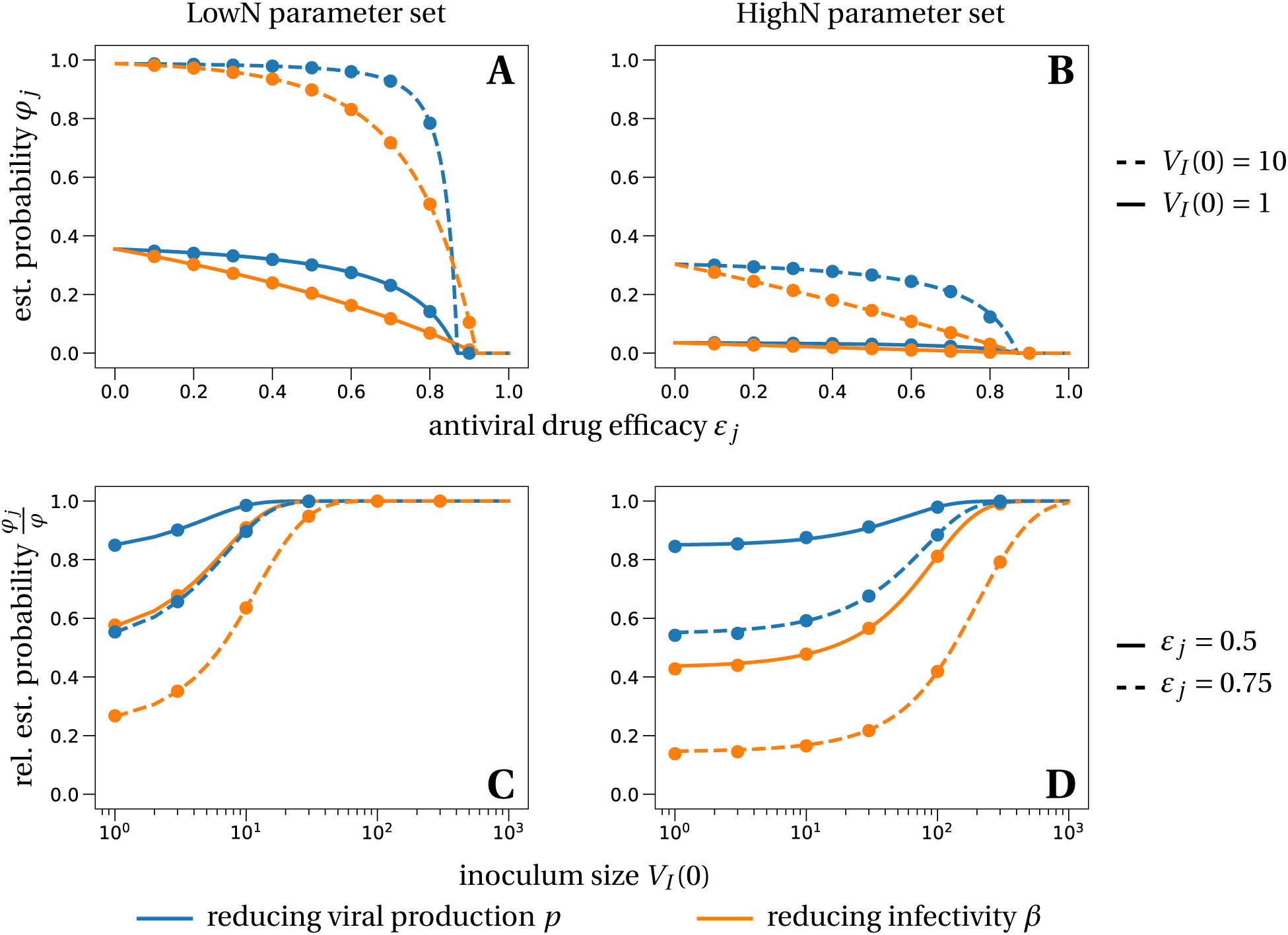
Establishment probability of the virus under different antiviral drugs, efficacies *ε* and various inoculum sizes *V*_0_. The lines in panels A and B correspond to the theoretical establishment probability under the assumption that target cell numbers are constant, for the two modes of action (reducing viral infectivity equivalent to increasing clearance, Eq. (5), in orange and reducing viral production equivalent to increasing cell death, Eq. (6), in blue). The lines in the bottom panels represent the relative probability of establishment normalized by the establishment probability in the absence of treatment from Eq. (4), i.e. *φ*_*j*_ /*φ*. Dots are averages from 100, 000 individual-based simulations of the within-host model described in system (3), in which target cell numbers are allowed to vary. Parameter values are given in Table 2.

#### 3.2.2 Antiviral increasing viral clearance

Antiviral drugs that increase the clearance rate *c* of extracellular virus particles reduce the average lifespan of a virus by a factor (1 −*ε*_*c*_). This changes the clearance parameter *c* by a factor 1/(1 −*ε*_*c*_).

With this definition of efficacy, we find that the reproductive number *R*_0_ is reduced by the same factor as obtained for a drug reducing infectivity: 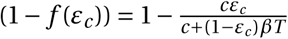. Therefore, the establishment probabilities take the same form, so that *φ*_*c*_ *= φ*_*β*_. Consequently, we will reduce our analysis to antiviral drugs that reduce viral infectivity, keeping in mind that results for the establishment probability are equally valid for drugs increasing viral clearance.

#### 3.2.3 Antiviral reducing viral production

Antiviral drugs reducing the viral production (parameter *p*) reduce the burst size *N* by a factor (1−*ε*_*p*_). The basic reproductive number *R*_0_ is reduced by the same factor. If (1 −*ε*_*p*_) × *R*_0_ ≥ 1, such drugs alter the establishment probability to:

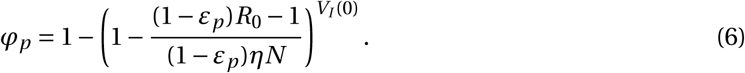

A reduction of 50% of the establishment probability compared to no treatment can be achieved with an efficacy of 85% (LowN parameter set, *V*_*I*_ (0) = 10). The efficacy needed is greater than that for antivirals targeting infectivity or viral clearance (81%) (see Fig. 2 panels A and C). Thus, for imperfect drugs that do not totally prevent establishment, drugs targeting infectivity (or clearance) are more efficient than those targeting viral production. This effect emerges from the stochastic dynamics and the reduction in viral production variance mentioned above: in the early phase, it is more important for the virus to infect many host cells than to ensure the production of a large number of virions. This insight might also affect the choice of antiviral drugs, depending on whether prophylaxis is taken pre- or post-exposure. In the case of pre-exposure, the scenario we mainly focus on and for which Eq. (4) was derived, we would recommend to prioritize drugs that increase extracellular viral clearance or reduce viral infectivity. A neutralizing monoclonal antibody such as LY-CoV555 could do both. On the other hand, if prophylactic treatment is started post-exposure, e.g. a couple of hours after a potential between-host transmission event, the likelihood is high that cells are already infected. If cells are infected, the initial condition of our analysis is changed and drugs reducing viral production such as a SARS-CoV-2 polymerase inhibitor or protease inhibitor are more efficient in preventing the establishment of the virus than drugs targeting extracellular viral processes (clearance and target cell infection) in the LowN parameter set, cf. Section S4 in the SI.

#### 3.2.4 Antiviral increasing infected cell death

Increasing the rate of death of infected cells *δ* by the factor 1/(1 −*ε*_*δ*_) reduces the average lifespan of an infected cell by a factor (1 −*ε*_*δ*_). This has the same effect on the burst size (and consequently on *R*_0_) as an antiviral drug reducing viral production, again due to our definition of efficacy. Therefore, the establishment probabilities are the same, *φ*_*p*_ *= φ*_*δ*_. In our analysis of establishment probabilities, we thus exclusively study antivirals affecting viral production.

#### 3.2.5 Critical efficacy

Above a critical treatment efficacy, the establishment of a viral infection is not possible. This is true for all modes of action and for high and low burst sizes (Fig. 2). The critical efficacy does not depend on the initial inoculum size. It is given by the condition that the drug-modified *R*_0_ equals 1, e.g. (1 −*ε*_*p*_)*R*_0_ = 1 for drugs reducing viral production *p*. This corresponds to the deterministic threshold value for the viral population to grow. Computing the critical efficacies for both modes of action with Eq. (5) and Eq. (6), we find:

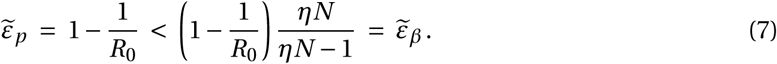

They differ for the two modes of action because reducing infectivity does not proportionally reduce *R*_0_ (Eq. (2)). Thus, drugs that reduce viral production result in a slightly lower critical efficacy, an effect that is small for a low burst size of infectious virions and not discernible with a high burst size of infectious virions (see intersections of the establishment probabilities with the x-axes in Fig. 2A and B). For example, in the HighN parameter set, we find a critical efficacy of 87% for both types of drugs.

In summary, in the range where drugs cannot totally prevent infection, drugs that target viral infectivity reduce the probability of establishment more strongly; drugs that reduce viral production can totally prevent infection at slightly lower efficacy, but this difference is extremely small when burst sizes (of infectious virions) are large.

### 3.3 Combination therapy

We analyze how the combination of two antiviral therapies could further impede establishment of the virus. We assume that two drugs that target different mechanisms of action lead to multiplicative effects on *R*_0_ (Bliss independence (Chou, 2006)). The establishment probability and critical efficacies for the two drugs can be computed in the same way as for single drug treatments.

For example, a combination of two drugs reducing viral production *p* and infectivity *β* changes the establishment probability to

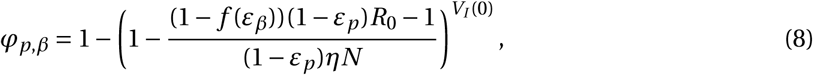

if (1 − *f* (*ε*_*β*_))(1 −*ε*_*p*_)*R*_0_ ≥ 1.

The corresponding critical pair of efficacies that prevent viral infection entirely can be computed as before by solving

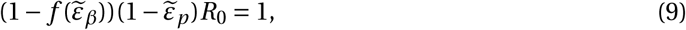

By the arguments from above, we can replace *ε*_*β*_ by *ε*_*c*_ and *ε*_*p*_ by *ε*_*δ*_ without changing the results. Similar calculations allow us to derive the analogous quantities if we combine drugs targeting the same mechanism of action, e.g. altering *p* and *δ* or *c* and *β* at the same time. Our analysis would also carry over to combination of drugs which target the same parameter if they interact multiplicatively. For example, two drugs reducing viral infectivity *β* with efficacies *ε*_*β*,1_ and *ε*_*β*,2_, respectively, would reduce *R*_0_ by the factor (1 − *f* (*ε*_*β*,1_))(1 − *f* (*ε*_*β*,2_)), if they act independently.

Using two drugs of limited efficacy in combination lead to large reductions in the establishment probability compared to the single drug or no treatment scenarios. For instance, two drugs with efficacies of 65% each may completely eliminate the risk of viral infection, depending on the combination used (LowN parameter set, *V*_*I*_ (0) = 1, Fig. 3). For comparison, a single drug with 65% efficacy can maximally reduce the establishment probability to ∼ 40% of the no-treatment establishment probability (see Fig. 2A). We also find that, compared to the single drug cases, the critical efficacy is significantly reduced in all combinations studied.

**Figure 3:**
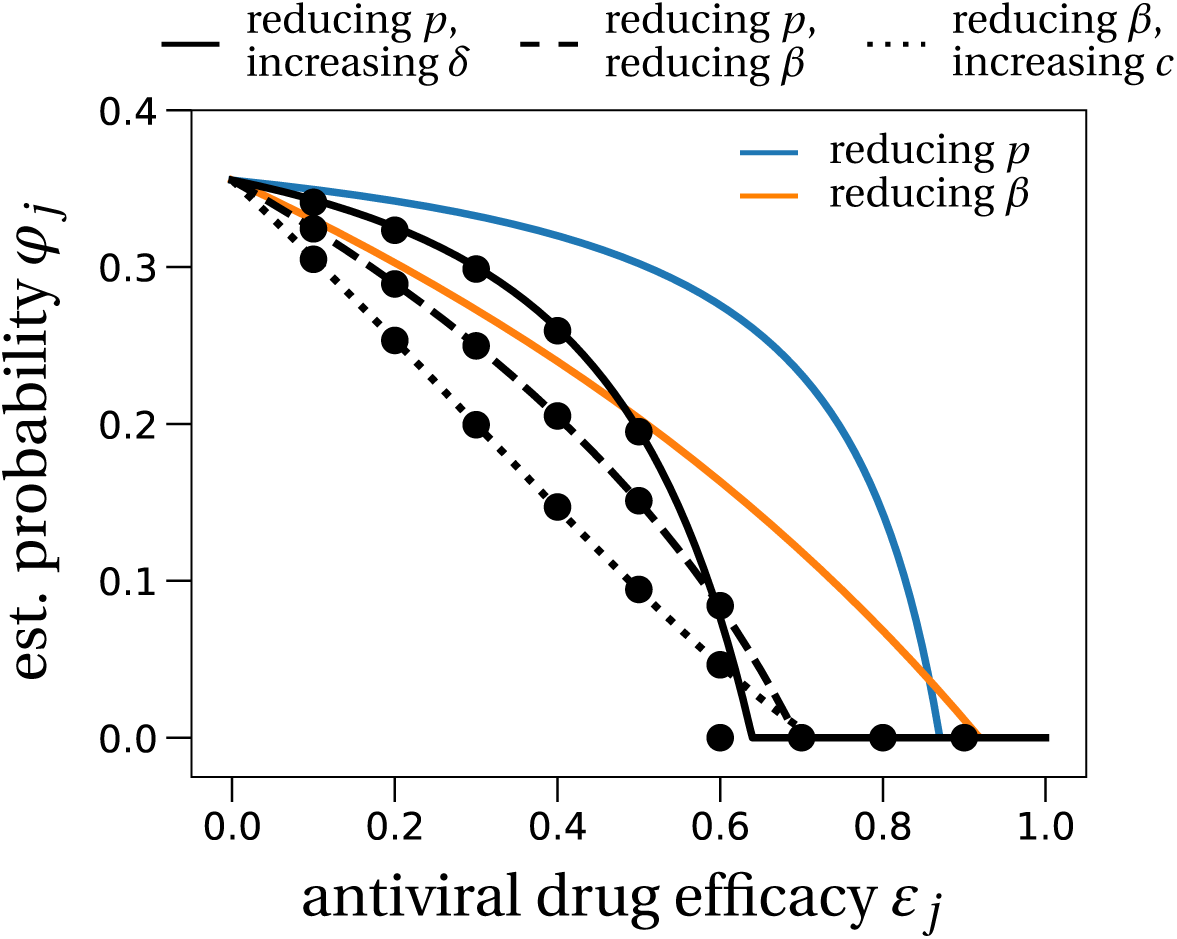
The effect of prophylactic combination therapy on the establishment probability. We compare different combination therapies (black lines) with the two single effect therapies (colored lines). The theoretical predictions for the combination therapies are variations of Eq. (8), adapted to the specific pair of modes of action considered. We assume that both modes of action are suppressed with the same efficacy, shown on the x-axis as *ε*_*j*_. Dots are averages from 100, 000 stochastic simulations using the LowN parameter set and *V*_*I*_ (0) = 1. In Section S5 in the SI, we study the effect of combination therapy in the HighN parameter set which overall leads to very similar results.

In our analysis, we assumed that the drugs act independently (Bliss independence). This assumption may lead to an over- or underestimation of the establishment probability in case of antagonistic or synergistic drug interactions, respectively. These interactions are difficult to anticipate but were observed for HIV treatments (Jilek et al., 2012).

### 3.4 Time to detectable viral load and extinction time

Lastly, we quantify the timescales of viral establishment and extinction of infectious virus particles. If the virus establishes, we ask whether therapy slows down its spread within the host and investigate how long it takes for the infection to reach the polymerase chain reaction (PCR) test detection threshold. Conversely, if the viral infection does not establish, we examine how long it takes for antiviral therapy to clear all infectious virus and infected cells, which we define as the extinction time. We study all four modes of action: drugs that increase either the infected cell death rate *δ* or viral clearance *c*, and drugs reducing either viral production *p* or the infectivity *β*.

#### 3.4.1 Time to detectable viral load

Even if antivirals are not efficacious enough to prevent establishment of the infection, could they still mitigate the infection? We study the effect of antiviral therapy on the time to reach a detectable viral load within the host. For example, the detection threshold in Young et al. (2020) is at 10^1.84^ copies per mL. Assuming that the upper respiratory tract has a volume of about 30 mL (Baccam et al., 2006b), this corresponds to approximately 2, 000 virus particles.

In our model without treatment, the viral population size reaches 2, 000 within one day (see the leftmost data point in Fig. 4). If establishment is likely, it is best to take antiviral drugs reducing the viral production *p* to delay the establishment of a viral infection as long as possible. This would reduce the peak viral load (Gonçalves et al., 2020; Goyal et al., 2020), which is presumably correlated with the severity of SARS-CoV-2 infection (Zheng et al., 2020). The time to reach a detectable viral load depends on the growth rate of the viral population, which is to the leading order 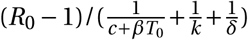 (see Section S5 in the SI for a derivation). The denominator is the average duration of a virus life cycle given by the sum of the phase when virions are in the medium, the eclipse phase of infected cells, and the phase during which infected cells produce virions until their death.

**Figure 4:**
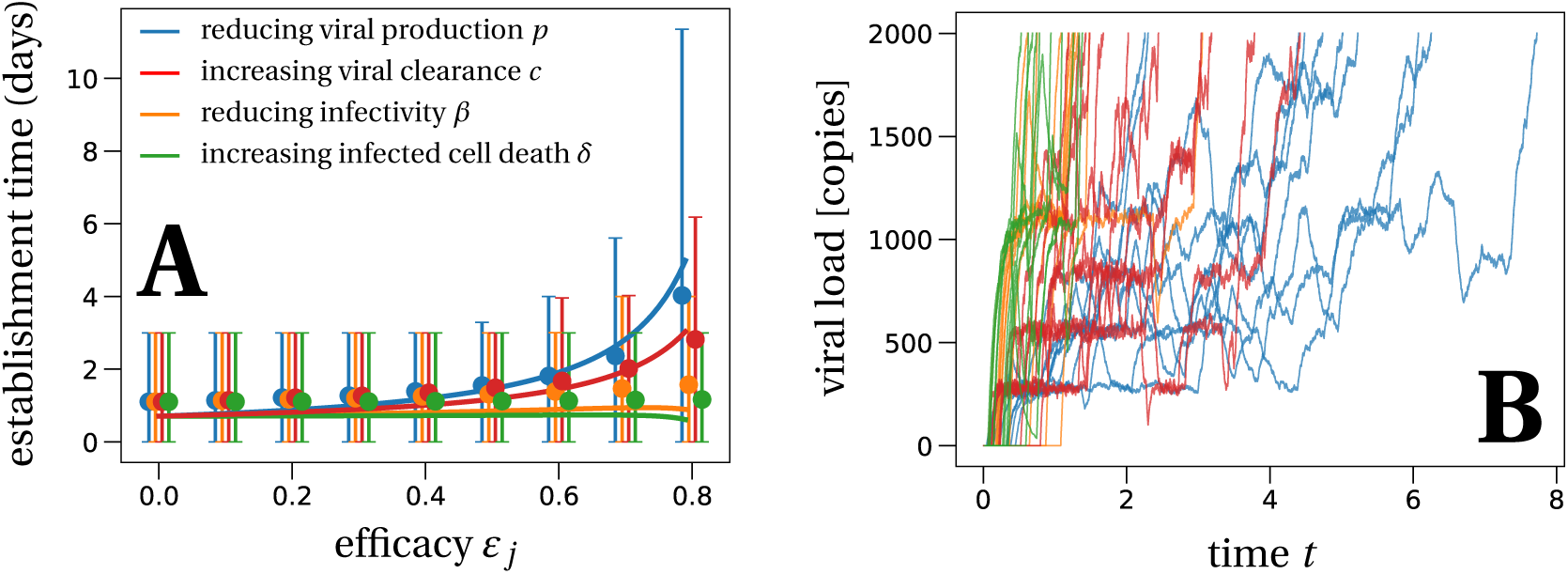
The mean time to reach a detectable viral load at the infection site. Panel A: Solid lines represent the theoretical prediction of the average time for the viral infection to reach 2, 000 virions (see Section S6 in the SI for details). We used the LowN parameter set to simulate 10, 000 stochastic simulations that reached a viral load of 2, 000 total virus particles when starting with an inoculum of *V*_*I*_ (0) = 1. Dots are the average times calculated from these simulations, error bars represent 90% of th simulated establishment times. Panel B: We plot 10 example trajectories that reach the detectable viral load for each of the four types of treatment (efficacy *ε*_*j*_ = 0.75). Under treatment that increases the infected cell death *δ* or reduces infectivity *β*, establishing trajectories reach the detectable viral load almost immediately. In contrast, drugs that directly affect the number of virus, i.e. clearance *c* or production *p*, allow for trajectories that fluctuate much more, explaining the larger average detection times and the larger variation of detection times for these scenarios.

Importantly, the time to reach a detectable viral load is the earliest time when a patient can be tested to determine if therapy succeeded or failed to prevent infection. That time can be increased up to 4 days for drugs inhibiting viral production *p* (blue line in Fig. 4), but there is significant variation with values ranging from smaller than one day to more than 10 days. Drugs reducing the infectivity *β* or increasing the infected cell death rate *δ* do not delay the establishment time. Drugs promoting viral clearance *c* increase the establishment time less than drugs decreasing the viral production rate *p*. As a brief explanation, when drugs target the infectivity or cell death, establishment occurs rapidly by full bursts of just two infected cells, which is enough to reach the detection threshold; when drugs target viral clearance or viral production, establishment may involve many more infected cells and occur slowly (SI Section S6.2).

#### 3.4.2 Extinction time of infectious virus particles

Given that the infection does not establish, extinction of the within-host population of infectious virus particles typically happens within a day (in the HighN parameter set) to up to a week (in the LowN parameter set) depending on the drug’s mode of action (Table 3). We find that antiviral drugs that either reduce viral infectivity *β* or increase infected cell death *δ* show comparably small extinction times (Table 3). The extinction time is useful to determine the number of days a potentially infected person should take antiviral medication post-exposure.

**Table 3:** Establishment probabilities, times to detection and extinction time statistics for various sets of antiviral treatment.

| $\varepsilon_j$ | Therapy | | LowN parameter set | | HighN parameter set | |
| --- | --- | --- | --- | --- | --- | --- |
| | | | $V_0 = 1$ | $V_0 = 10$ | $V_0 = 1$ | $V_0 = 10$ |
| 0 | no treatment | $\varphi$ | 36% | 99% | 4% | 30% |
| | | $T_{\text{detect}}$ | 1 (0.5, 1.5) | 0.5 (0, 0.5) | 0.5 (0, 1) | 0 (0, 0.5) |
| | | $T_{\text{ext}}$ | 0 (0, 0) | 1 (0, 1.5) | 0 (0, 0) | 0.5 (0, 0.5) |
| 0.75 | reducing $p$ | $\varphi$ | 20% | 89% | 2% | 18% |
| | | $T_{\text{detect}}$ | 4 (2, 9) | 2 (0.5, 6.5) | 0.5 (0, 1) | 0.5 (0, 1) |
| | | $T_{\text{ext}}$ | 0 (0, 2) | 2.5 (1, 6) | 0 (0, 0) | 0.5 (0, 2) |
| | increasing $\delta$ | $\varphi$ | 20% | 89% | 2% | 18% |
| | | $T_{\text{detect}}$ | 1 (0.5, 2) | 0.5 (0, 1) | 0.5 (0, 1) | 0 (0, 0.5) |
| | | $T_{\text{ext}}$ | 0 (0, 1.5) | 1.5 (1, 3) | 0 (0, 0) | 0.5 (0, 1.5) |
| | reducing $\beta$ | $\varphi$ | 9% | 63% | 1% | 5% |
| | | $T_{\text{detect}}$ | 1 (0.5, 2.5) | 0.5 (0.5, 2) | 0.5 (0, 1) | 0.5 (0, 1) |
| | | $T_{\text{ext}}$ | 0 (0, 0.5) | 0.5 (0, 2.5) | 0 (0, 0) | 0.5 (0, 0.5) |
| | increasing $c$ | $\varphi$ | 9% | 63% | 1% | 5% |
| | | $T_{\text{detect}}$ | 2.5 (1.5, 5.5) | 2 (1, 5) | 0 (0, 0.5) | 0 (0, 0.5) |
| | | $T_{\text{ext}}$ | 0 (0, 0) | 0 (0, 2) | 0 (0, 0) | 0 (0, 0) |
| 0.9 | reducing $p$ | $\varphi$ | 0% | 0% | 0% | 0% |
| | | $T_{\text{detect}}$ | – | – | – | – |
| | | $T_{\text{ext}}$ | 0 (0, 5) | 7 (2.5, 19) | 0 (0, 0.5) | 0.5 (0, 5) |
| | increasing $\delta$ | $\varphi$ | 0% | 0% | 0% | 0% |
| | | $T_{\text{detect}}$ | – | – | – | – |
| | | $T_{\text{ext}}$ | 0 (0, 2) | 2.5 (1, 5) | 0.5 (0, 1) | 0.5 (0, 2) |
| | reducing $\beta$ | $\varphi$ | 1% | 11% | 0% | 0% |
| | | $T_{\text{detect}}$ | 1.5 (0.5, 3.5) | 1 (0.5, 3) | – | – |
| | | $T_{\text{ext}}$ | 0 (0, 0.5) | 0.5 (0, 6) | 0 (0, 0) | 0.5 (0, 0.5) |
| | increasing $c$ | $\varphi$ | 1% | 11% | 0% | 0% |
| | | $T_{\text{detect}}$ | 12 (5.5, 29) | 12 (5, 28) | – | – |
| | | $T_{\text{ext}}$ | 0 (0, 0) | 0 (0, 30) | 0 (0, 0) | 0 (0, 0) |

The first value in each cell gives the establishment probability, the second value denotes the median time to detection (days), the numbers in brackets are the 10 and 90-percentiles of the time to detection distribution (days), and the last line of numbers gives the median time to extinction (days), conditioned on non-establishment of the infection, with the 10 and 90-percentiles in brackets. The detection threshold is set to 2,000 virus particles. All times are rounded to half-day values if below 5 days, and to days if above. Missing values, denoted by dashes, are explained by the viral population not establishing; values above 30 days are set to 30. All results are estimated from 100,000 stochastic simulations for the establishment probability and 10,000 stochastic trajectories for the extinction and establishment times.

## 4 Discussion

We have investigated the effect of prophylaxis with antiviral treatments including monoclonal antibodies on the viral dynamics of SARS-CoV-2. Using a stochastic model of within-host SARS-CoV-2 dynamics whose structure and parameters are informed by recent data (Gonçalves et al., 2020; Kim et al., 2020), we showed that in principle a combination of two drugs each with efficacy between 60% and 70% will almost certainly prevent infection (Fig. 3). For single drug treatment, we find that even intermediate efficacies can block infection, most efficiently with drugs reducing infectivity *β*, or otherwise delay the within-host establishment of the viral infection for drugs reducing viral production *p* or increasing viral clearance *c* (Fig. 4). More generally, our stochastic model for the early phase of virus establishment within a host could be used to study the impact of prophylactic treatment on viral infections whose dynamics can be captured by the deterministic model in Eq. (1).

This model makes several important assumptions. First, it encompasses a simplified version of the innate immune response. Effects of this type of immune reaction are embedded in the parameter values of the model. For example, an early innate response, if not effectively subverted by the virus, might put some target cells into an antiviral state where they are refractory to infection, thus effectively reducing *β* (Pawelek et al., 2012), or it could reduce the viral production rate *p* (Baccam et al., 2006a). We neglect a potential adaptive immune response against the virus because we are interested in the early stages of the infection, before the immune system develops a specific response to the viral infection. A specific immune response may in later stages enhance the ability of the body to eliminate the virus. Thus, the estimates of the drug efficacies needed to prevent establishment of infection are conservative and in reality may be overestimates. Further, even if the drugs being used do not have efficacies high enough to prevent infection on their own, they can lengthen the time needed to establish infection and hence allow time for the immune response to develop and assist in the clearance of the virus. Lastly, we focus on the early phase of the infection in the upper respiratory tract, and neglect other compartments that may be more favorable to viral multiplication. For example, the number of virions in the sputum is (on average) 10 to 100 fold higher than in throat swabs (Pan et al., 2020). The upper respiratory tract may allow a small amount of virus to enter the lower respiratory tract. It would be interesting in future work to explore the impact of this spatial structure on viral dynamics and establishment probability.

Our results on critical efficacy, shown in Figs. 2 and 3, do not depend on the viral inoculum size and are very similar for low and high burst sizes. However, they strongly depend on the intrahost basic reproductive number which we estimated at *R*_0_ = 7.69. This basic reproductive number was estimated from time series of viral load in nasopharyngeal swabs in 13 infected patients (Young et al., 2020; Gonçalves et al., 2020) and is consistent with the mean peak viral load observed in multiple studies (Table 1). Still, there is substantial inter-individual heterogeneity in incubation time, observed peak viral timing and load (He et al., 2020). A shorter time to the viral load peak or a higher viral load peak would result in higher estimates of *R*_0_, see for instance Fig. 1B. Yet, our qualitative findings on the effectiveness of prophylactic therapy remain valid under these variations of *R*_0_. Of course, the quantitative predictions, which depend on *R*_0_, change. Considering the current uncertainty in the basic reproductive number and burst size, we developed an interactive application to compute and visualize the establishment probability and deterministic dynamics as a function of parameters. This application can be used to update our results as our knowledge of intrahost dynamics and treatment efficacies progresses (it can be accessed by following the instructions on github.com/pczuppon/virus_establishment/tree/master/shiny).

The critical efficacy above which infection is entirely prevented is the efficacy at which the intrahost basic reproductive number, adjusted to the antiviral drug under consideration, passes below 1. The value of this critical efficacy could readily be obtained in a deterministic model. Yet, our stochastic framework gives several new additional insights into the probability of establishment. Importantly, below the critical efficacy, viral establishment is not certain. The establishment probability increases with the size of the initial inoculum (Fig. 2). The number of infectious virions of seasonal coronavirus in droplets and aerosol particles exhaled during 30 minutes could be in the range of 1 to 10 (Leung et al., 2020). Assuming this to be the range of the inoculum of infectious virus particles, in most cases the establishment of a viral infection is not ensured even with low-efficacy drugs. For efficacies below the critical efficacy, drugs reducing infectivity or increasing viral clearance reduce the establishment probability the most. Examples for this type of drug include monoclonal neutralizing antibodies that recently have shown promising results for treatment and prophylaxis of SARS-CoV-2 (Baum et al., 2020). In contrast, drugs reducing viral production need to be close to critical efficacy to cause a marked reduction on the probability of establishment (Figs. 2 and 3). Several studies are underway to assess the prophylactic potential of repurposed drugs blocking viral production, such as lopinavir, favipiraivr or remdesivir, but there is no clear demonstration that these drugs can achieve clinically relevant antiviral efficacy (Fragkou et al., 2020; WHO Solidarity trial consortium et al., 2020; Beigel et al., 2020).

Similar theoretical results have been obtained for HIV antiviral prophylactic treatments (Duwal et al., 2019). If initially there is one infectious HIV particle, drugs that target viral production within cells are less successful in inhibiting infection than drugs that reduce viral infection of target cells, cf. Fig. 2A in Duwal et al. (2019). However, if the virus has already infected a cell, the difference between the two drug types vanishes, i.e., both modes of action equally reduce the establishment of an infection (Figs. 2B, 2C in Duwal et al. (2019)). In contrast, with our model we find that if there is initially one infected cell, establishment of a viral infection is suppressed more strongly by drugs that reduce viral production than by those reducing infection of target cells (Section S4 in the SI). This difference most likely arises due to the different burst sizes of infectious virus particles assumed in the two models. Here, we assume that the burst size is around 20 infectious virus particles, computed by *η*× *N*. In contrast, the HIV model studied in Duwal et al. (2019) assumes a burst size of 670. Indeed, increasing the burst size in our model, the HighN parameter set, recovers the result found in Duwal et al. (2019), i.e., the two different drug types affect the establishment probability equally.

Lastly, we observe that given that extinction occurs the time to extinction is largely independent of the drug’s mode of action and typically occurs within a day (see Table 3). In contrast, we find a relatively strong dependence of the time to detection of an infection on the mode of action of the antiviral drug. The time to detection also strongly depends on the burst size which varies substantially depending on the assumed fraction of infectious virus particles produced, *η*. For example, a lower fraction than considered here in the main text will result in a higher burst size for a fixed value of *R*_0_ (Section S7.2 in the SI) and consequently in a lower time to detection. If the delay between exposure and therapy, as well as the efficacy of the available drugs, are such that establishment of the viral infection is almost certain, antiviral drugs that reduce viral production (parameter *p*) will slow down the exponential growth and flatten the within-host epidemic curve the most (Fig. 4). Repurposed antiviral drugs reducing viral production were recently proposed as good drug candidates against SARS-CoV-2 (Gordon et al., 2020). This prolonged period at low viral loads could give the immune system the necessary time to activate a specific response to the virus and develop temporary host-immunity against SARS-CoV-2. This might be especially important in groups that are frequently exposed to the virus, e.g. health care workers. Still, since reducing the infection probability itself is the primary goal, drugs reducing the infectivity of virus (parameters *β* and *c*) should be favored over drugs reducing viral production (parameters *p* and *δ*) because of their stronger effect on the establishment probability (Fig. 2).

## Conclusion

Clinical trials are underway to test the efficacy of several antiviral drugs (Harrison, 2020; Li and Clercq, 2020; Fragkou et al., 2020; Sheahan et al., 2020; Maisonnasse et al., 2020), either as a curative treatment or as a prevention. The efficacy of repurposed drugs is in a 20-70% range (Gonçalves et al., 2020), but better antiviral drugs might be available soon. Thus, our prediction that a drug efficacy needs to be greater than 90% to prevent infection establishment may be within reach in a near future. An interactive tool has been made available to update this prediction with refined parameter estimates that will come from large dataset obtained in the different target populations where prophylaxis may be relevant (such as health care workers or high-risk individuals). Given the current knowledge of SARS-CoV-2 viral dynamics, our model predicts that prophylactic antiviral therapy can block (or at least delay) a viral infection, could be administered to people at risk such as health care workers, and alleviate the burden on the healthcare systems caused by the SARS-CoV-2 pandemic.

## Methods

### Simulations

The individual based simulations are coded in C++ using the standard stochastic simulation algorithm for the reactions described in system (3).

Estimates for the establishment probabilities, depicted by dots in the subsequent figures, are averages of 100, 000 independent runs. Establishment was considered successful when the population size of infectious virions was at least 500. Estimates for the time to reach a detectable viral load are obtained from 10, 000 simulations where the sum of infectious and non-infectious virus particles exceeded 2, 000 copies.

The code and the data to generate the figures are available at: github.com/pczuppon/virus_ establishment.

## Data Availability

The codes and data to generate the figures are publicly available on a github repository as stated in the manuscript.

## Acknowledgments

P.C. has received funding from the European Union’s Horizon 2020 research and innovation program under the Marie Sklodowska-Curie grant agreement PolyPath 844369. This project was partially funded by the ANR TheraCov (ANR-20-COVI-0018) and the BMGF foundation under grant agreement INV-017335. O.T. and F.B. received funding from Grant Equipe Fondation pour la Recherche Médicale EQU201903007848. F.B. received funding from the Centre National de la Recherche Scientifique (Momentum grant). F.D. received funding from the national research agency (ANR) through the grant ANR-19-CE45-0009-01. Portions of this work were done under the auspices of the US Department of Energy under Contract 89233218CNA000001 (A.S.P.). This work was also supported by the Los Alamos National Laboratory LDRD program (A.S.P.). We are grateful to the INRA MIGALE bioinformatics facility (MIGALE, INRA, 2018. Migale bioinformatics Facility, doi: 10.15454/1.5572390655343293E12) for providing computational resources.

## Notes

### Competing Interest Statement

The authors have declared no competing interest.

### Funding Statement

P.C. has received funding from the European Union's Horizon 2020 research and innovation program under the Marie Skłodowska-Curie grant agreement PolyPath 844369. This project was partially funded by the ANR TheraCov (ANR-20-COVI-0018) and the BMGF foundation under grant agreement INV- 017335. O.T. and F.B. received funding from Grant Equipe Fondation pour la Recherche Médicale EQU201903007848. F.B. received funding from the Centre National de la Recherche Scientifique (Momentum grant). F.D. received funding from the national research agency (ANR) through the grant ANR-19-CE45-0009-01.
Portions of this work were done under the auspices of the US Department of Energy under Contract 89233218CNA000001 (A.S.P.). This work was also supported by the Los Alamos National Laboratory LDRD program (A.S.P.). We are grateful to the INRA MIGALE bioinformatics facility (MIGALE, INRA, 2018. Migale bioinformatics Facility, doi: 10.15454/1.5572390655343293E12) for providing computational resources.

### Author Declarations

No experimental data was generated.

### Summary of Updates

updated manuscript and data

## References

Baccam, P., Beauchemin, C., Macken, C. A., Hayden, F. G., & Perelson, A. S. (2006a). Kinetics of Influenza A virus infection in humans. Journal of Virology, 80(15):7590–7599. doi: 10.1128/JVI.01623-05.

Baccam, P., Beauchemin, C., Macken, C. A., Hayden, F. G., & Perelson, A. S. (2006b). Kinetics of Influenza A Virus Infection in Humans. Journal of Virology, 80(15):7590–7599. doi: 10.1128/JVI.01623-05.

Baeten, J. M., Donnell, D., Ndase, P., Mugo, N. R., Campbell, J. D., Wangisi, J., Tappero, J. W., Bukusi, E. A., Cohen, C. R., Katabira, E., Ronald, A., Tumwesigye, E., Were, E., Fife, K. H., Kiarie, J., Farquhar, C., John-Stewart, G., Kakia, A., Odoyo, J., Mucunguzi, A., Nakku-Joloba, E., Twesigye, R., Ngure, K., Apaka, C., Tamooh, H., Gabona, F., Mujugira, A., Panteleeff, D., Thomas, K. K., Kidoguchi, L., Krows, M., Revall, J., Morrison, S., Haugen, H., Emmanuel-Ogier, M., Ondrejcek, L., Coombs, R. W., Frenkel, L., Hendrix, C., Bumpus, N. N., Bangsberg, D., Haberer, J. E., Stevens, W. S., Lingappa, J. R., & Celum, C. (2012). Antiretroviral prophylaxis for HIV prevention in heterosexual men and women. New England Journal of Medicine, 367(5):399–410. doi: 10.1056/NEJMoa1108524.

Bar-On, Y. M., Flamholz, A., Phillips, R., & Milo, R. (2020). SARS-CoV-2 (COVID-19) by the numbers. eLife, 9. doi: 10.7554/eLife.57309.

Baum, A., Copin, R., Ajithdoss, D., Zhou, A., Lanza, K., Negron, N., Ni, M., Wei, Y., Atwal, G. S., Oyejide, A., Goez-Gazi, Y., Dutton, J., Clemmons, E., Staples, H. M., Bartley, C., Klaffke, B., Alfson, K., Gazi, M., Gonzales, O., Dick, E., Carrion, R., Pessaint, L., Porto, M., Cook, A., Brown, R., Ali, V., Greenhouse, J., Taylor, T., Andersen, H., Lewis, M. G., Stahl, N., Murphy, A. J., Yancopoulos, G. D., & Kyratsous, C. A. (2020). REGN-COV2 antibody cocktail prevents and treats SARS-CoV-2 infection in rhesus macaques and hamsters. medRxiv. doi: 10.1101/2020.08.02.233320.

Beigel, J. H., Tomashek, K. M., Dodd, L. E., Mehta, A. K., Zingman, B. S., Kalil, A. C., Hohmann, E., Chu, H. Y., Luetkemeyer, A., Kline, S., de Castilla, D. L., Finberg, R. W., Dierberg, K., Tapson, V., Hsieh, L., Patterson, T. F., Paredes, R., Sweeney, D. A., Short, W. R., Touloumi, G., Lye, D. C., Ohmagari, N., don Oh, M., Ruiz-Palacios, G. M., Benfield, T., Fätkenheuer, G., Kortepeter, M. G., Atmar, R. L., Creech, C. B., Lundgren, J., Babiker, A. G., Pett, S., Neaton, J. D., Burgess, T. H., Bonnett, T., Green, M., Makowski, M., Osinusi, A., Nayak, S., & Lane, H. C. (2020). Remdesivir for the treatment of Covid-19 — final report. New England Journal of Medicine, 383(19):1813–1826. doi: 10.1056/nejmoa2007764.

Bi, Q., Wu, Y., Mei, S., Ye, C., Zou, X., Zhang, Z., Liu, X., Wei, L., Truelove, S. A., Zhang, T., Gao, W., Cheng, C., Tang, X., Wu, X., Wu, Y., Sun, B., Huang, S., Sun, Y., Zhang, J., Ma, T., Lessler, J., & Feng, T. (2020). Epidemiology and transmission of COVID-19 in 391 cases and 1286 of their close contacts in Shenzhen, China: a retrospective cohort study. The Lancet Infectious Diseases. doi: 10.1016/s1473-3099(20)30287-5.

Cereda, D., Tirani, M., Rovida, F., Demicheli, V., Ajelli, M., Poletti, P., Trentini, F., Guzzetta, G., Marziano, V., Barone, A., Magoni, M., Deandrea, S., Diurno, G., Lombardo, M., Faccini, M., Pan, A., Bruno, R., Parliani, E., Grasselli, G., Piatti, A., Gramegna, A., Baldanti, F., Melegaro, A., & Merler, S. (2020). The early phase of the COVID-19 outbreak in Lombardy, Italy. 2003.09320 [q-bio].

Chen, P., Nirula, A., Heller, B., Gottlieb, R. L., Boscia, J., Morris, J., Huhn, G., Cardona, J., Mocherla, B., Stosor, V., Shawa, I., Adams, A. C., Naarden, J. V., Custer, K. L., Shen, L., Durante, M., Oakley, G., Schade, A. E., Sabo, J., Patel, D. R., Klekotka, P., & Skovronsky, D. M. (2020). SARS-CoV-2 neutralizing antibody LY-CoV555 in outpatients with Covid-19. New England Journal of Medicine. doi: 10.1056/nejmoa2029849.

Chinazzi, M., Davis, J. T., Ajelli, M., Gioannini, C., Litvinova, M., Merler, S., Piontti, A. P. y., Mu, K., Rossi, L., Sun, K., Viboud, C., Xiong, X., Yu, H., Halloran, M. E., Longini, I. M., & Vespignani, A. (2020). The effect of travel restrictions on the spread of the 2019 novel coronavirus (COVID-19) outbreak. Science. doi: 10.1126/science.aba9757.

Chou, T.-C. (2006). Theoretical basis, experimental design, and computerized simulation of synergism and antagonism in drug combination studies. Pharmacological Reviews, 58(3):621–681. doi: 10.1124/pr.58.3.10.

Conway, J. M., Konrad, B. P., & Coombs, D. (2013). Stochastic analysis of pre- and postexposure prophylaxis against HIV infection. SIAM Journal on Applied Mathematics, 73(2):904–928. doi: 10.1137/120876800.

Dong, E., Du, H., & Gardner, L. (2020). An interactive web-based dashboard to track COVID-19 in real time. The Lancet Infectious Diseases, 20(5):533–534. doi: https://doi.org/10.1016/S1473-3099(20)30120-1.

Duwal, S., Dickinson, L., Khoo, S., & von Kleist, M. (2019). Mechanistic framework predicts drug-class specific utility of antiretrovirals for HIV prophylaxis. PLOS Computational Biology, 15(1):e1006740. doi: 10.1371/journal.pcbi.1006740.

Ferguson, N. M., Laydon, D., Nedjati-Gilani, G., Imai, N., Ainslie, K., Baguelin, M., Bhatia, S., Boonyasiri, A., CucunubÃ¡ , Z., Cuomo-Dannenburg, G., Dighe, A., Fu, H., Gaythorpe, K., Thompson, H., Verity, R., Volz, E., Wang, H., Wang, Y., Walker, P. G., Walters, C., Winskill, P., Whittaker, C., Donnelly, C. A., Riley, S., & Ghani, A. C. (2020). Impact of non-pharmaceutical interventions (NPIs) to reduce COVID-19 mortality and healthcare demand. Imperial College London - COVID-19 reports, Report 9.

Ferretti, L., Wymant, C., Kendall, M., Zhao, L., Nurtay, A., Abeler-Dörner, L., Parker, M., Bonsall, D., & Fraser, C. (2020). Quantifying SARS-CoV-2 transmission suggests epidemic control with digital contact tracing. Science, 368(6491). doi: 10.1126/science.abb6936.

Fragkou, P., Belhadi, D., Peiffer-Smadja, N., Moschopoulos, C., Lescure, F.-X., Janocha, H., Karofylakis, E., Yazdanpanah, Y., Mentré, F., Skevaki, C., Laouénan, C., & Tsiodras, S. (2020). Review of trials currently testing treatment and prevention of COVID-19. Clinical Microbiology and Infection, 26 (8):988–998. doi: 10.1016/j.cmi.2020.05.019.

Gonçalves, A., Bertrand, J., Ke, R., Comets, E., de Lamballerie, X., Malvy, D., Pizzorno, A., Terrier, O., Rosa Calatrava, M., Mentré, F., Smith, P., Perelson, A. S., & Guedj, J. (2020). Timing of antiviral treatment initiation is critical to reduce SARS-CoV-2 viral load. CPT: Pharmacometrics & Systems Pharmacology, 9(9). doi: 10.1002/psp4.12543.

Gordon, D. E., Jang, G. M., Bouhaddou, M., Xu, J., Obernier, K., White, K. M., O’Meara, M. J., Rezelj, V. V., Guo, J. Z., Swaney, D. L., Tummino, T. A., Hüttenhain, R., Kaake, R. M., Richards, A. L., Tutuncuoglu, B., Foussard, H., Batra, J., Haas, K., Modak, M., Kim, M., Haas, P., Polacco, B. J., Braberg, H., Fabius, J. M., Eckhardt, M., Soucheray, M., Bennett, M. J., Cakir, M., McGregor, M. J., Li, Q., Meyer, B., Roesch, F., Vallet, T., Kain, A. M., Miorin, L., Moreno, E., Naing, Z. Z. C., Zhou, Y., Peng, S., Shi, Y., Zhang, Z., Shen, W., Kirby, I. T., Melnyk, J. E., Chorba, J. S., Lou, K., Dai, S. A., Barrio-Hernandez, I., Memon, D., Hernandez-Armenta, C., Lyu, J., Mathy, C. J. P., Perica, T., Pilla, K. B., Ganesan, S. J., Saltzberg, D. J., Rakesh, R., Liu, X., Rosenthal, S. B., Calviello, L., Venkataramanan, S., Liboy-Lugo, J., Lin, Y., Huang, X.-P., Liu, Y., Wankowicz, S. A., Bohn, M., Safari, M., Ugur, F. S., Koh, C., Savar, N. S., Tran, Q. D., Shengjuler, D., Fletcher, S. J., O’Neal, M. C., Cai, Y., Chang, J. C. J., Broadhurst, D. J., Klippsten, S., Sharp, P. P., Wenzell, N. A., Kuzuoglu-Ozturk, D., Wang, H.-Y., Trenker, R., Young, J. M., Cavero, D. A., Hiatt, J., Roth, T. L., Rathore, U., Subramanian, A., Noack, J., Hubert, M., Stroud, R. M., Frankel, A. D., Rosenberg, O. S., Verba, K. A., Agard, D. A., Ott, M., Emerman, M., Jura, N., von Zastrow, M., Verdin, E., Ashworth, A., Schwartz, O., d’Enfert, C., Mukherjee, S., Jacobson, M., Malik, H. S., Fujimori, D. G., Ideker, T., Craik, C. S., Floor, S. N., Fraser, J. S., Gross, J. D., Sali, A., Roth, B. L., Ruggero, D., Taunton, J., Kortemme, T., Beltrao, P., Vignuzzi, M., García-Sastre, A., Shokat, K. M., Shoichet, B. K., & Krogan, N. J. (2020). A SARS-CoV-2 protein interaction map reveals targets for drug repurposing. Nature, 583(7816):459–468. doi: 10.1038/s41586-020-2286-9.

Goyal, A., Cardozo-Ojeda, E. F., & Schiffer, J. T. (2020). Potency and timing of antiviral therapy as determinants of duration of SARS CoV-2 shedding and intensity of inflammatory response. medRxiv. doi: 10.1101/2020.04.10.20061325.

Harrison, C. (2020). Coronavirus puts drug repurposing on the fast track. Nature Biotechnology. doi: 10.1038/d41587-020-00003-1.

Hauser, A., Counotte, M. J., Margossian, C. C., Konstantinoudis, G., Low, N., Althaus, C. L., & Riou, J. (2020). Estimation of SARS-CoV-2 mortality during the early stages of an epidemic: a modelling study in Hubei, China and northern Italy. medRxiv. doi: 10.1101/2020.03.04.20031104.

He, X., Lau, E. H. Y., Wu, P., Deng, X., Wang, J., Hao, X., Lau, Y. C., Wong, J. Y., Guan, Y., Tan, X., Mo, X., Chen, Y., Liao, B., Chen, W., Hu, F., Zhang, Q., Zhong, M., Wu, Y., Zhao, L., Zhang, F., Cowling, B. J., Li, F., & Leung, G. M. (2020). Temporal dynamics in viral shedding and transmissibility of COVID-19. Nature Medicine, 26(5):672–675. doi: 10.1038/s41591-020-0869-5.

Hirano, N., Fujiwara, K., & Matumoto, M. (1976). Mouse Hepatitis Virus (MHV-2). Japanese Journal of Microbiology, 20(3):219–225. doi: 10.1111/j.1348-0421.1976.tb00978.x.

Hoffmann, R. M., Mele, S., Cheung, A., Larcombe-Young, D., Bucaite, G., Sachouli, E., Zlatareva, I., Morad, H. O. J., Marlow, R., McDonnell, J. M., Figini, M., Lacy, K. E., Tutt, A. J. N., Spicer, J. F., Thurston, D. E., Karagiannis, S. N., & Crescioli, S. (2020). Rapid conjugation of antibodies to toxins to select candidates for the development of anticancer antibody-drug conjugates (ADCs). Scientific Reports, 10(1). doi: 10.1038/s41598-020-65860-x.

Hurford, A., Cownden, D., & Day, T. (2010). Next-generation tools for evolutionary invasion analyses. Journal of the Royal Society Interface, 7(45). doi: 10.1098/rsif.2009.0448.

Igarashi, T., Brown, C., Azadegan, A., Haigwood, N., Dimitrov, D., Martin, M. A., & Shibata, R. (1999). Human immunodeficiency virus type 1 neutralizing antibodies accelerate clearance of cell–free virions from blood plasma. Nature Medicine, 5(2):211–216. doi: 10.1038/5576.

Jiang, S., Hillyer, C., & Du, L. (2020). Neutralizing antibodies against SARS-CoV-2 and other human coronaviruses. Trends in Immunology, 41(5):355–359. doi: https://doi.org/10.1016/j.it.2020.03.007.

Jilek, B. L., Zarr, M., Sampah, M. E., Rabi, S. A., Bullen, C. K., Lai, J., Shen, L., & Siliciano, R. F. (2012). A quantitative basis for antiretroviral therapy for HIV-1 infection. Nature Medicine, 18(3):446–451. doi: 10.1038/nm.2649.

Jones, T. C., Mühlemann, B., Veith, T., Biele, G., Zuchowski, M., Hoffmann, J., Stein, A., Edelmann, A., Corman, V. M., & Drosten, C. (2020). An analysis of SARS-CoV-2 viral load by patient age. medRxiv.

Ke, R., Zitzmann, C., Ribeiro, R. M., & Perelson, A. S. (2020). Kinetics of SARS-CoV-2 infection in the human upper and lower respiratory tracts and their relationship with infectiousness. medRxiv. doi: 10.1101/2020.09.25.20201772.

Kim, K. S., Ejima, K., Ito, Y., Iwanami, S., Ohashi, H., Koizumi, Y., Asai, Y., Nakaoka, S., Watashi, K., Thompson, R. N., & Iwami, S. (2020). Modelling SARS-CoV-2 dynamics: Implications for therapy. medRxiv. doi: 10.1101/2020.03.23.20040493.

Kissler, S. M., Fauver, J. R., Mack, C., Tai, C., Shiue, K. Y., Kalinich, C. C., Jednak, S., Ott, I. M., Vogels, C. B., Wohlgemuth, J., Weisberger, J., DiFiori, J., Anderson, D. J., Mancell, J., Ho, D. D., Grubaugh, N. D., & Grad, Y. H. (October 2020). Viral dynamics of SARS-CoV-2 infection and the predictive value of repeat testing. medRxiv. doi: 10.1101/2020.10.21.20217042. URL https://doi.org/10.1101/2020.10.21.20217042.

Knoops, K., Kikkert, M., Van Den Worm, S. H., Zevenhoven-Dobbe, J. C., Van Der Meer, Y., Koster, A. J., Mommaas, A. M., & Snijder, E. J. (2008). SARS-coronavirus replication is supported by a reticulovesicular network of modified endoplasmic reticulum. PLoS Biol, 6(9):e226.

Lai, S., Bogoch, I., Ruktanonchai, N., Watts, A., Lu, X., Yang, W., Yu, H., Khan, K., & Tatem, A. J. (2020). Assessing spread risk of Wuhan novel coronavirus within and beyond China, January-April 2020: a travel network-based modelling study. medRxiv. doi: 10.1101/2020.02.04.20020479.

Leung, N. H. L., Chu, D. K. W., Shiu, E. Y. C., Chan, K.-H., McDevitt, J. J., Hau, B. J. P., Yen, H.-L., Li, Y., Ip, D. K. M., Peiris, J. S. M., Seto, W.-H., Leung, G. M., Milton, D. K., & Cowling, B. J. (2020). Respiratory virus shedding in exhaled breath and efficacy of face masks. Nature Medicine, 26(5): 676–680. doi: 10.1038/s41591-020-0843-2.

Li, G. & Clercq, E. D. (2020). Therapeutic options for the 2019 novel coronavirus (2019-nCoV). Nature Reviews Drug Discovery, 19(3). doi: 10.1038/d41573-020-00016-0.

Li, Q., Guan, X., Wu, P., Wang, X., Zhou, L., Tong, Y., Ren, R., Leung, K. S., Lau, E. H., Wong, J. Y., Xing, X., Xiang, N., Wu, Y., Li, C., Chen, Q., Li, D., Liu, T., Zhao, J., Liu, M., Tu, W., Chen, C., Jin, L., Yang, R., Wang, Q., Zhou, S., Wang, R., Liu, H., Luo, Y., Liu, Y., Shao, G., Li, H., Tao, Z., Yang, Y., Deng, Z., Liu, B., Ma, Z., Zhang, Y., Shi, G., Lam, T. T., Wu, J. T., Gao, G. F., Cowling, B. J., Yang, B., Leung, G. M., & Feng, Z. (2020). Early transmission dynamics in Wuhan, China, of novel coronavirus–infected pneumonia. New England Journal of Medicine, 382(13):1199–1207. doi: 10.1056/NEJMoa2001316.

Maisonnasse, P., Guedj, J., Contreras, V., Behillil, S., Solas, C., Marlin, R., Naninck, T., Pizzorno, A., Lemaitre, J., Gonçalves, A., Kahlaoui, N., Terrier, O., Fang, R. H. T., Enouf, V., Dereuddre-Bosquet, N., Brisebarre, A., Touret, F., Chapon, C., Hoen, B., Lina, B., Calatrava, M. R., van der Werf, S., de Lamballerie, X., & Grand, R. L. (2020). Hydroxychloroquine use against SARS-CoV-2 infection in non-human primates. Nature, 585(7826):584–587. doi: 10.1038/s41586-020-2558-4.

Mermin, J., Ekwaru, J. P., Liechty, C. A., Were, W., Downing, R., Ransom, R., Weidle, P., Lule, J., Coutinho, A., & Solberg, P. (2006). Effect of co-trimoxazole prophylaxis, antiretroviral therapy, and insecticide-treated bednets on the frequency of malaria in HIV-1-infected adults in Uganda: a prospective cohort study. The Lancet, 367(9518):1256–1261. doi: 10.1016/S0140-6736(06)68541-3.

Muniz-Rodriguez, K., Chowell, G., Cheung, C.-H., Jia, D., Lai, P.-Y., Lee, Y., Liu, M., Ofori, S. K., Roosa, K. M., Simonsen, L., Viboud, C., & Fung, I. C.-H. (2020). Doubling time of the COVID-19 epidemic by Province, China. Emerging Infectious Diseases, 26(8). doi: 10.3201/eid2608.200219.

Munster, V. J., Feldmann, F., Williamson, B. N., van Doremalen, N., Pérez-Pérez, L., Schulz, J., Meade-White, K., Okumura, A., Callison, J., Brumbaugh, B., Avanzato, V. A., Rosenke, R., Hanley, P. W., Saturday, G., Scott, D., Fischer, E. R., & de Wit, E. (2020). Respiratory disease in rhesus macaques inoculated with SARS-CoV-2. Nature. doi: 10.1038/s41586-020-2324-7.

National Institute of Allergy and Infectious Diseases (NIAID). SARS-CoV-2: Images and B-roll related to the novel coronavirus (SARS-CoV-2, also known as 2019-nCoV) that causes COVID-19., (2020 (accessed November 5, 2020)). URL https://www.flickr.com/photos/niaid/albums/72157712914621487/with/49531042907/.

Pagliano, P., Piazza, O., De Caro, F., Ascione, T., & Filippelli, A. (2020). Is hydroxychloroquine a possible postexposure prophylaxis drug to limit the transmission to healthcare workers exposed to coronavirus disease 2019? Clinical Infectious Diseases. doi: 10.1093/cid/ciaa320.

Pan, Y., Zhang, D., Yang, P., Poon, L. L. M., & Wang, Q. (2020). Viral load of SARS-CoV-2 in clinical samples. The Lancet Infectious Diseases, 20(4):411–412. doi: 10.1016/S1473-3099(20)30113-4.

Pawelek, K. A., Huynh, G. T., Quinlivan, M., Cullinane, A., Rong, L., & Perelson, A. S. (06 2012). Modeling within-host dynamics of influenza virus infection including immune responses. PLOS Computational Biology, 8(6):1–13. doi: 10.1371/journal.pcbi.1002588.

Pearson, J. E., Krapivsky, P., & Perelson, A. S. (02 2011). Stochastic theory of early viral infection: Continuous versus burst production of virions. PLOS Computational Biology, 7(2):1–17. doi: 10.1371/journal.pcbi.1001058.

Robb, J. A. & Bond, C. W. Coronaviridae. In Fraenkel-Conrat, H. & Wagner, R. R., editors, Comprehensive Virology: Newly Characterized Vertebrate Viruses, Comprehensive Virology, pages 193–247. Springer US, Boston, MA, (1979). doi: 10.1007/978-1-4684-3563-4_3.

Salje, H., Tran Kiem, C., Lefrancq, N., Courtejoie, N., Bosetti, P., Paireau, J., Andronico, A., Hozé, N., Richet, J., Dubost, C.-L., Le Strat, Y., Lessler, J., Levy-Bruhl, D., Fontanet, A., Opatowski, L., Boelle, P.-Y., & Cauchemez, S. (2020). Estimating the burden of sars-cov-2 in france. Science. doi: 10.1126/science.abc3517.

Sheahan, T. P., Sims, A. C., Zhou, S., Graham, R. L., Pruijssers, A. J., Agostini, M. L., Leist, S. R., Schäfer, A., Dinnon, K. H., Stevens, L. J., Chappell, J. D., Lu, X., Hughes, T. M., George, A. S., Hill, C. S., Montgomery, S. A., Brown, A. J., Bluemling, G. R., Natchus, M. G., Saindane, M., Kolykhalov, A. A., Painter, G., Harcourt, J., Tamin, A., Thornburg, N. J., Swanstrom, R., Denison, M. R., & Baric, R. S. (2020). An orally bioavailable broad-spectrum antiviral inhibits SARS-CoV-2 in human airway epithelial cell cultures and multiple coronaviruses in mice. Science Translational Medicine, 12 (541). doi: 10.1126/scitranslmed.abb5883.

Spinelli, F. R., Ceccarelli, F., Di Franco, M., & Conti, F. (2020). To consider or not antimalarials as a prophylactic intervention in the SARS-CoV-2 (COVID-19) pandemic. Annals of the Rheumatic Diseases, 79(5):666–667. doi: 10.1136/annrheumdis-2020-217367.

Stertz, S., Reichelt, M., Spiegel, M., Kuri, T., Martínez-Sobrido, L., García-Sastre, A., Weber, F., & Kochs, G. (2007). The intracellular sites of early replication and budding of SARS-coronavirus. Virology, 361(2):304–315.

Tindale, L., Coombe, M., Stockdale, J. E., Garlock, E., Lau, W. Y. V., Saraswat, M., Lee, Y.-H. B., Zhang, L., Chen, D., Wallinga, J., & Colijn, C. (2020). Transmission interval estimates suggest pre-symptomatic spread of COVID-19. medRxiv. doi: 10.1101/2020.03.03.20029983.

To, K. K.-W., Tsang, O. T.-Y., Leung, W.-S., Tam, A. R., Wu, T.-C., Lung, D. C., Yip, C. C.-Y., Cai, J.-P., Chan, J. M.-C., Chik, T. S.-H., Lau, D. P.-L., Choi, C. Y.-C., Chen, L.-L., Chan, W.-M., Chan, K.-H., Ip, J. D., Ng, A. C.-K., Poon, R. W.-S., Luo, C.-T., Cheng, V. C.-C., Chan, J. F.-W., Hung, I. F.-N., Chen, Z., Chen, H., & Yuen, K.-Y. (May 2020). Temporal profiles of viral load in posterior oropharyngeal saliva samples and serum antibody responses during infection by SARS-CoV-2: an observational cohort study. The Lancet Infectious Diseases, 20(5):565–574. doi: 10.1016/s1473-3099(20)30196-1.

Tubiana, S., Burdet, C., Houhou, N., Thy, M., Manchon, P., Blanquart, F., Charpentier, C., Guedj, J., Alavoine, L., Behillil, S., et al. (2020). High-risk exposure without personal protective equipment and infection with sars-cov-2 in healthcare workers: results of the cov-contact prospective cohort. medRxiv.

US National Library of Medicine. ClinicalTrials.gov is a database of privately and publicly funded clinical studies conducted around the world, (2020 (accessed November 12, 2020)). URL https://www.clinicaltrials.gov/ct2/home.

Verity, R., Okell, L. C., Dorigatti, I., Winskill, P., Whittaker, C., Imai, N., Cuomo-Dannenburg, G., Thompson, H., Walker, P. G. T., Fu, H., Dighe, A., Griffin, J. T., Baguelin, M., Bhatia, S., Boonyasiri, A., Cori, A., Cucunubá, Z., FitzJohn, R., Gaythorpe, K., Green, W., Hamlet, A., Hinsley, W., Laydon, D., Nedjati-Gilani, G., Riley, S., van Elsland, S., Volz, E., Wang, H., Wang, Y., Xi, X., Donnelly, C. A., Ghani, A. C., & Ferguson, N. M. (2020). Estimates of the severity of coronavirus disease 2019: a model-based analysis. The Lancet Infectious Diseases, 20(6):669–677. doi: 10.1016/s1473-3099(20)30243-7.

WHO Solidarity trial consortium, Pan, H., Peto, R., Karim, Q. A., Alejandria, M., Henao-Restrepo, A. M., García, C. H., Kieny, M.-P., Malekzadeh, R., Murthy, S., Preziosi, M.-P., Reddy, S., Periago, M. R., Sathiyamoorthy, V., Røttingen, J.-A., & and, S. S. (2020). Repurposed antiviral drugs for COVID-19 –interim WHO SOLIDARITY trial results. medRxiv. doi: 10.1101/2020.10.15.20209817.

Wölfel, R., Corman, V. M., Guggemos, W., Seilmaier, M., Zange, S., Müller, M. A., Niemeyer, D., Jones, T. C., Vollmar, P., Rothe, C., Hoelscher, M., Bleicker, T., Brünink, S., Schneider, J., Ehmann, R., Zwirglmaier, K., Drosten, C., & Wendtner, C. (2020). Virological assessment of hospitalized patients with COVID-2019. Nature, 581(7809):465–469. doi: 10.1038/s41586-020-2196-x.

Wu, J. T., Leung, K., Bushman, M., Kishore, N., Niehus, R., Salazar, P. M. d., Cowling, B. J., Lipsitch, M., & Leung, G. M. (2020). Estimating clinical severity of COVID-19 from the transmission dynamics in Wuhan, China. Nature Medicine. doi: 10.1038/s41591-020-0822-7.

Young, B. E., Ong, S. W. X., Kalimuddin, S., Low, J. G., Tan, S. Y., Loh, J., Ng, O.-T., Marimuthu, K., Ang, L. W., Mak, T. M., Lau, S. K., Anderson, D. E., Chan, K. S., Tan, T. Y., Ng, T. Y., Cui, L., Said, Z., Kurupatham, L., Chen, M. I.-C., Chan, M., Vasoo, S., Wang, L.-F., Tan, B. H., Lin, R. T. P., Lee, V. J. M., Leo, Y.-S., & and, D. C. L. (2020). Epidemiologic features and clinical course of patients infected with SARS-CoV-2 in Singapore. JAMA, 323(15):1488. doi: 10.1001/jama.2020.3204.

Zheng, S., Fan, J., Yu, F., Feng, B., Lou, B., Zou, Q., Xie, G., Lin, S., Wang, R., Yang, X., Chen, W., Wang, Q., Zhang, D., Liu, Y., Gong, R., Ma, Z., Lu, S., Xiao, Y., Gu, Y., Zhang, J., Yao, H., Xu, K., Lu, X., Wei, G., Zhou, J., Fang, Q., Cai, H., Qiu, Y., Sheng, J., Chen, Y., & Liang, T. (2020). Viral load dynamics and disease severity in patients infected with SARS-CoV-2 in Zhejiang province, China, January-March 2020: retrospective cohort study. BMJ, 369. doi: 10.1136/bmj.m1443.

Zhu, N., Zhang, D., Wang, W., Li, X., Yang, B., Song, J., Zhao, X., Huang, B., Shi, W., Lu, R., Niu, P., Zhan, F., Ma, X., Wang, D., Xu, W., Wu, G., Gao, G. F., & Tan, W. (2020). A novel coronavirus from patients with pneumonia in China, 2019. New England Journal of Medicine, 382(8):727–733. doi: 10.1056/NEJMoa2001017.

