## Supplementary Information for "Success of prophylactic antiviral therapy for SARS-CoV-2: predicted critical efficacies and impact of different drug-specific mechanisms of action"

#### Contents

**S1 Continuous virus production model** **2**
S1.1 Connection to a burst model . . . . . . . . . . . . . . . . . . . . . . . . . . . . . 2

**S2 Burst model** **4**
S2.1 Establishment probability . . . . . . . . . . . . . . . . . . . . . . . . . . . . . . 4

**S3 Comparison of the continuous-production and burst model** **6**

**S4 Establishment probability when starting with a single infected cell** **8**

**S5 Combination therapy in the HighN parameter set** **10**

**S6 Time to detectable viral load** **11**

### S1 Continuous virus production model

We recall the model from the main text. Infectious and non-infectious virus particles are denoted by $V_I$ and $V_{NI}$, respectively, target cells by $T$, infected cells in the eclipse phase by $I_1$ and infected cells producing the virus by $I_2$. We use a previously studied within-host model of virus production (Pearson et al., 2011; Conway et al., 2013). The underlying individual based reactions are the following:

$$\begin{aligned}
&V_I + T \xrightarrow{\beta} I_1, && \text{infection of target cells,}\\
&I_1 \xrightarrow{k} I_2, && \text{end of eclipse phase,}\\
&I_2 \xrightarrow{\delta} \varnothing, && \text{infected cell death,}\\
&I_2 \xrightarrow{\eta p} I_2 + V_I, && \text{infectious virus production,}\\
&I_2 \xrightarrow{(1-\eta)p} I_2 + V_{NI}, && \text{non-infectious virus production,}\\
&V_I, V_{NI} \xrightarrow{c} \varnothing, && \text{virus clearance.}
\end{aligned} \tag{S1}$$

Since we model the early state within-host dynamics of a viral infection, we can assume that the number of infectious virus particles, $V_I$, is low so that the number of target cells is not strongly affected by transformation to infected cells, i.e. $T(t) \approx T(0) = T_0$. Then, the first reaction can be rewritten as

$$V_I \xrightarrow{\beta T_0} I_1\,. \tag{S2}$$

Using standard techniques to derive a set of ordinary differential equations from this set of reactions (e.g. Anderson and Kurtz, 2011), we find the system given in eq. (1) in the main text. Note, that in the main text we use capital letters to denote densities while here the capital letters refer to the actual numbers of cells and virus particles.

#### S1.1 Connection to a burst model

Since the individual based model is built on stochastic interactions of cells and virions, the number of virions produced by an infected cell is a random variable. Assuming that all virions are released at a single time, typically at cell death, the number of released virions, the burst size, follows a geometric distribution (Hataye et al., 2019). This can be seen by the following reasoning: the life-time of an infected cell is exponentially distributed with mean $1/\delta$ and during this time there is a continuous production of virions at rate $p$. This production, assuming that it is a Markovian process, is described by a Poisson process (see Anderson and Kurtz (2011) for the general theory of modeling chemical reactions). The probability of the burst size, denoted by $N$, to be of size $j$ is then given by the following calculation:

$$\begin{aligned}\mathbb{P}(N=j) &= \int_0^\infty \underbrace{\frac{(pt)^j}{j!}e^{-pt}}_{j \text{ virions produced until time } t} \underbrace{\delta e^{-\delta t}}_{\text{cell still alive at time } t} dt \\ &= \frac{p^j \delta}{j!}\int_0^\infty t^j e^{-(p+\delta)t} dt \\ &= \left(\frac{p}{p+\delta}\right)^j \frac{\delta}{p+\delta}.\end{aligned} \tag{S3}$$

This is the distribution of a geometrically distributed random variable with success probability $p/(p+\delta)$. Intuitively, the infected cell has undergone $j+1$ steps, where the initial $j$ steps resulted in the production of a virus (term $(p/(p+\delta))^j$) and the $(j+1)$-th step was its death (term $\delta/(p+\delta)$). The mean of this geometric distribution is $p/\delta$. The continuous-production model can therefore be seen as equivalent to a burst size model with a burst size $N$ having a geometric distribution with mean $p/\delta$.

#### S2 Burst model

The continuous-production model is more likely to be relevant for SARS-CoV-2 (Park et al., 2020), and was therefore chosen in the main text. Here we examine how a burst model would affect our findings. In a burst model, we assume that virus is produced in an infected cell but is only released to the environment upon cell death. The number of virus particles released is therefore a random number which we again denote by $N$.

In the corresponding reactions in Eq. (S1), we need to replace the virus production and cell death lines by

$$I_2 \xrightarrow{\delta} \eta N V_I + (1-\eta) N V_{NI}. \tag{S4}$$

In order to be consistent with the continuous-production model, we set the mean of the burst size to $p/\delta$.

In the following we assume that the overall burst size $N$ is Poisson-distributed. There are two reasons for this choice: (i) it is analytically relatively easy to handle, and (ii) it represents the other end of the spectrum of negative-binomially distributed burst sizes when compared to the continuous-production model which is equivalent to a geometrically-distributed burst size and thus providing an upper bound for the establishment probability of a viral infection under different forms of virus release from infected cells. A negative binomial distribution is defined by a success probability $q$ and a dispersion parameter $r$. The mean is given by $qr/(1-q)$. It relates to the geometric distribution by setting $r = 1$ and to the Poisson distribution by letting $r \to \infty$. The probabilities of establishment for the continuous-production model and the Poisson-distributed burst size model represent the two extremes of negative-binomially distributed burst size models with dispersion parameter $r \in (1, \infty)$. This holds because the establishment probability can be computed by the probability generating function (Haccou et al., 2005) which is continuous and monotone in the dispersion parameter $r$. It is given by

$$g(z) = \left(\frac{1-q}{1-qz}\right)^r, \tag{S5}$$

where $z$ is an auxiliary variable.

##### S2.1 Establishment probability

We compute the establishment probability of the virus in the burst size model. A key ingredient is the offspring distribution of a single virus particle. The offspring distribution is given by a zero-inflated Poisson distribution:

$$\begin{aligned} \mathbb{P}(0 \text{ infectious virus offspring}) &= \underbrace{\frac{c}{c+\beta T}}_{\text{no cell infected}} + \underbrace{\frac{\beta T}{c+\beta T} e^{-\eta N}}_{\text{infected cell with 0 virions produced}}, \\ \mathbb{P}(j \text{ infectious virus offspring}) &= \frac{\beta T}{c+\beta T} \frac{(\eta N)^j}{j!} e^{-\eta N}, \quad \text{for } j \in \{1,2,3,\ldots\}. \end{aligned} \tag{S6}$$

Note, that we are only considering infectious virus particles here because non-infectious virus particles do not affect the future virus dynamics.

The life cycle of a virus (conditioned on infecting a cell) is given by a three step process: cell infection, eclipse phase and virus production within an infected cell. Ignoring this time delay which is irrelevant if we just consider the establishment probability, the virus population can be modeled by a discrete time branching process. At each time, all infectious virions alive at the time step before produce a random number of (infectious) virions according to the offspring distribution given in eq. (S6).

The extinction probability of a time-discrete branching process, when starting with one infectious virus particle, is given by the non-trivial fixed point of the probability generating function of the offspring distribution, i.e. the fixed point in the interval $(0,1)$ (Haccou et al., 2005). The probability generating function is given by

$$g(z) = \mathbb{E}[z^{\eta N}] = \frac{c}{c+\beta T} + \frac{\beta T}{c+\beta T} e^{\eta N(z-1)}, \tag{S7}$$

where $z$ is an auxiliary variable and $\mathbb{E}$ denotes the expectation of the random burst size of infectious virions $\eta N$. The fixed point of this function is given as

$$z^* = \frac{c}{c+\beta T} - \frac{W\left(-\eta N \exp\left(-\eta N \frac{\beta T}{c+\beta T}\right)\right)}{\eta N}, \tag{S8}$$

where $W(x)$ is the Lambert-function (sometimes also called the product logarithm). It is defined for $x \geq -\exp(-1)$. For values below this threshold, we need to solve eq. (S7) numerically. In fact, when plotting the establishment probability in Fig. S1 below, we solve eq. (S7) numerically because the approximation of the Lambert-W function $W(x)$ is inaccurate for negative $x$, especially when close to $-\exp(-1)$.

The establishment probability, denoted $\varphi$, is then given by

$$\mathbb{P}(\text{virus survives}) = \varphi = 1 - \min(1, z^*)^{V_I(0)}, \tag{S9}$$

where $V_I(0)$ is the initial number of infectious virions. For alternative derivations of this result see also Pearson et al. (2011) and Conway et al. (2013).

#### S3 Comparison of the continuous-production and burst model

We compare the establishment probability from the burst model described above with that obtained in the continuous-production model. Redrawing the first row of Fig. 1 from the main text and comparing it with the corresponding graphs obtained from the burst model, we do not see any qualitative difference between the two models, cf. Fig. S1. As outlined in Section S2, the two studied models can be seen as the extreme values of a model continuum. By varying the dispersal parameter $r$ of the negative binomial distribution, one can explore the entire continuum between the geometrically distributed burst size (which is equivalent to the continuous-production model) and the Poisson-distributed burst size model. Therefore, it seems safe to say that the exact mechanism by which we implement virus production in the model will only result in (minor) quantitative differences on the probability of virus establishment.

| **Parameter set** | $\eta p$ $[d^{-1}]$ | $T_0$ **[cells]** | $\eta N$ **[virions]** | $R_0$ **[cells]** |
|---|---|---|---|---|
| Low burst size (LowN) | 11.2 | $4 \times 10^4$ | 18.8 | 7.69 |
| High burst size (HighN) | 112 | $4 \times 10^3$ | 188 | 7.69 |

Table S1: **Model parameters used in the main text and for the simulations in Fig. S1.** The remaining parameters are not changed between the simulations and are set to: $k = 5\ d^{-1}$, $\delta = 0.595\ d^{-1}$, $c = 10\ d^{-1}$, $\beta = c\delta R_0/(T_0(\eta p - \delta R_0))\ d^{-1}$, $\eta = 0.001$.

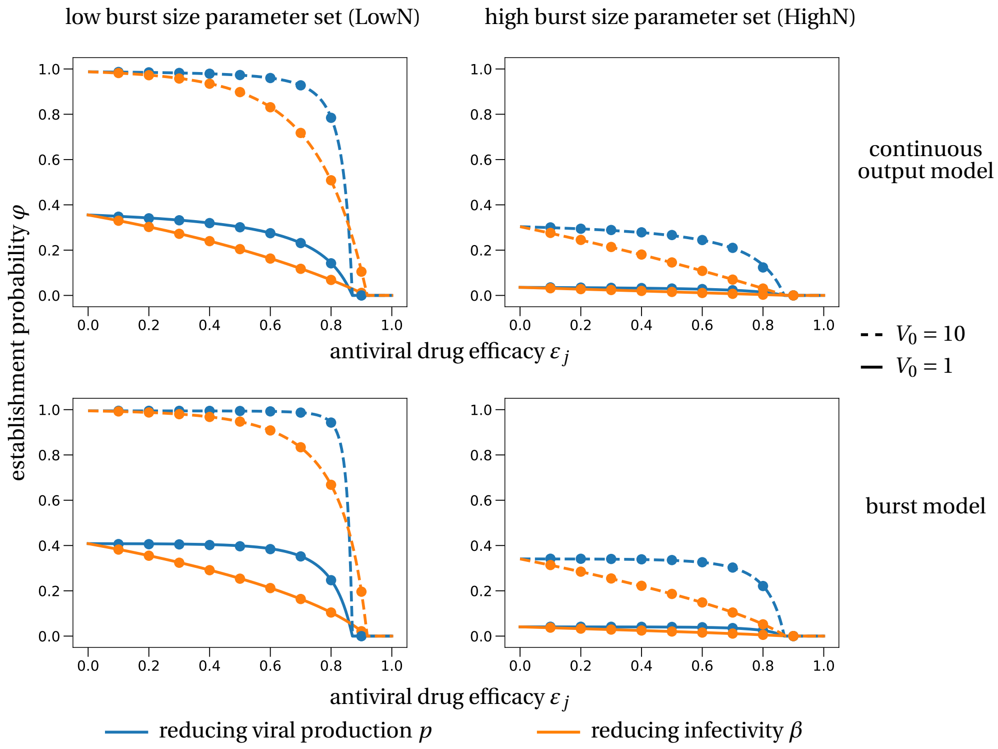

Figure S1: **Comparison of establishment probabilities in the continuous-production and burst model.** The first row is the same as the first row in Fig. 2 in the main text. The second row corresponds to the burst model. Theoretical approximations of the establishment probability for the burst model are obtained from Eq. (S9) adapted to the different scenarios.

### S4 Establishment probability when starting with a single infected cell

In this section, we investigate how the establishment probability changes if treatment is started when there is already an infected cell within the host. This situation might be more realistic to post-exposure treatment where infectious virus from the initial inoculum might have already infected a target cell (if the virus was not cleared). Instead of starting with a viral inoculum, we thus need to consider the situation where an infectious cell is already producing virus (but has not yet produced an infectious virus particle). The reasoning for computing the establishment probability is then as follows: we combine the establishment probability with initially $j$ infectious virus particles with the probability for this infected cell to produce $j$ infectious virus particles. As we have seen in Section S1.1 the number of infectious virus particles produced by an infectious cell is geometrically distributed with success parameter $\delta/(\delta+\eta p)$. Therefore, the establishment probability when starting with an infected cell, denoted by $\varphi_I$, is given by

$$\begin{aligned}
\psi &= \sum_{j=1}^{\infty} \underbrace{\left(\frac{\eta p}{\eta p+\delta}\right)^j \left(\frac{\delta}{\eta p+\delta}\right)}_{j \text{ infectious virus particles}} \underbrace{\left(1-(1-\varphi)^j\right)}_{\text{est. prob. for } j \text{ inf. virions}} \\
&= \left(\frac{\delta}{\eta p+\delta}\right) \sum_{j=1}^{\infty} \left(\frac{\eta p}{\eta p+\delta}\right)^j \left(1-\left(1-\frac{R_0-1}{\eta N}\right)^j\right) \\
&= \frac{p(R_0-1)}{\delta N + p(R_0-1)} \\
&= 1-\frac{1}{R_0}.
\end{aligned} \tag{S10}$$

This result has also been derived in Duwal et al. (2019) for a similar model in the context of HIV prophylaxis.

In our high burst size parameter set, there is no visible difference between treatment with a drug reducing productivity $p$ and a drug reducing viral infectivity $\beta$ (Fig. S2b). However, for the low burst size parameter set, in contrast to what we found in the main text when initializing the system with a viral inoculum, now drugs reducing the infectivity $p$ (blue) stronger reduce the establishment probability than drugs reducing the infectivity $\beta$ (orange), cf. Fig. S2a. This is explained by the order in which the drugs act: while a drug reducing viral production can immediately lower the chances for a further virus propagation, drugs reducing infectivity need to ‘wait’ for their targets, the extra-cellular virus, to arrive.

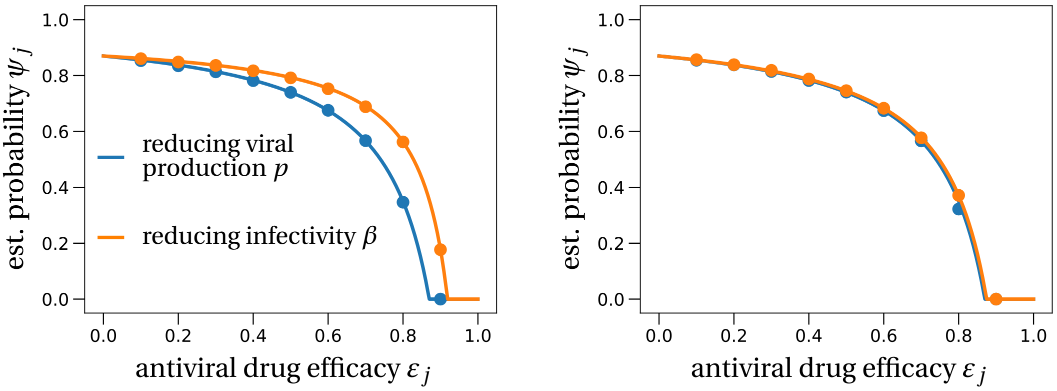

(a) LowN parameter set

(b) HighN parameter set

Figure S2: **Establishment probability when starting with a single infected cell.** We compare the theoretical prediction (solid lines) of Eq. (S10), adjusted for an antiviral drug affecting either virus productivity or virus infectivity, with stochastic simulations in the (a) LowN and (b) HighN parameter set. In the theoretical derivation of the results, target cells are fixed to their initial values. In the stochastic simulations, this number is allowed to decrease after cell infection. Averages of 10,000 realizations are depicted as dots. In contrast to the finding in the main text, in the LowN parameter set drugs reducing viral production $p$ reduce the establishment probability stronger than antivirals reducing infectivity $\beta$. This difference becomes negligible in the HighN parameter set.

#### S5 Combination therapy in the HighN parameter set

We investigate the effect of combination therapy in the high burst size parameter set (Table S1). We find that the overall shape of the curves do not change compared to the LowN parameter set. A higher burst size decreases the establishment probability of the virus. If we compare Fig. S3(b) with Fig. 3 in the main text, we see that a ten-fold increase of the initial inoculum in the HighN parameter set ($V_0 = 10$) gives similar quantitative results as the LowN parameter set with $V_0 = 1$. This can be attributed to our ten-fold increase of the burst size when deriving the HighN parameter set from the LowN parameter set.

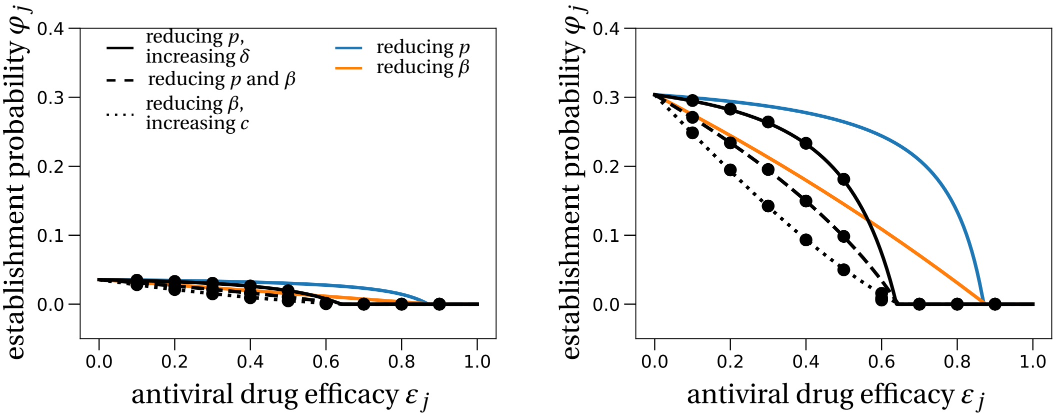

(a) $V_0 = 1$ (b) $V_0 = 10$

Figure S3: **Combination therapy in the HighN parameter set.** We plot the establishment probability of different combination therapies as was done in Fig. 3 in the main text. Dots are averages from 100,000 stochastic simulations obtained using the HighN parameter set with (a) $V_0 = 1$ or (b) $V_0 = 10$.

### S6 Time to detectable viral load

In this section, we study the mean time to reach a certain amount of viral load at the infection site within the host. We approximate this time using a mixture of deterministic and stochastic arguments. Classical branching processes typically have two possible outcomes: either the process goes extinct or grows indefinitely (Haccou et al., 2005). The deterministic model is captured by the mean of such a branching process, i.e. it takes into account both possible outcomes. Therefore, if we condition the branching process on survival, the deterministic model will typically underestimate the actual size of the corresponding branching process (Desai and Fisher, 2007). One can correct this error by rescaling the deterministic process by the probability of survival. In our specific setting this means that the total number of virus particles at any time $t$, $V(t) = V_I(t) + V_{NI}(t)$, can be estimated as follows:

$$\begin{aligned} &\mathbb{E}[V(t)] = \varphi(t)\mathbb{E}[V(t); V(t) > 0] + (1-\varphi(t))\underbrace{\mathbb{E}[V(t); V(t) = 0]}_{=0} \\ \iff \quad &\mathbb{E}[V(t); V(t) > 0] = \frac{\mathbb{E}[V(t)]}{\varphi(t)}, \end{aligned} \tag{S11}$$

where $V(t)$ denotes the random variable for the number of virus particles at time $t$, $\varphi(t)$ the survival probability of the branching process until time $t$ and $\mathbb{E}[V(t); V(t) > 0]$ the expectation of $V(t)$ for a surviving trajectory until time $t$.

To compute the time for the viral load to reach a certain threshold we set $\varphi(t) = \varphi$. In other words, we approximate the survival of the branching process until time $t$ by the total establishment probability expressed in eq. (4) in the main text. This is a good approximation if the 'typical' time $t$ to reach the threshold is large enough, so that $\varphi(t)$ is already close to the limit survival probability $\varphi$. The other term on the right-hand side in eq. (S11), the mean of the stochastic process $\mathbb{E}[V(t)]$, can be approximated by the deterministic model of the within-host model defined in eq. (1) in the main text.

As explained in the main text, we set the threshold viral load 2,000 virions (Fig. 4 in the main text). The mean time to reach this threshold value is then approximated by the time when the size $2{,}000 \times \varphi$ is reached in the deterministic model.

#### S6.1 Growth rate of the viral population to leading order

The exponential growth rate of the deterministic model described in eq. (1) in the main text is given by the leading eigenvalue of the system when evaluated at the origin, i.e. at zero virions Bonhoeffer et al. (1997). For efficacies close to the critical efficacy, the eigenvalue is small and can therefore be approximated by the root of a linear equation instead of a higher order polynomial. This approximation yields $\frac{R_0-1}{\frac{1}{c+\beta T_0}+\frac{1}{k}+\frac{1}{\delta}}$ as the leading eigenvalue. A *Mathematica* notebook showing this calculation is deposited at: `gitlab.com/pczuppon/virus_establishment`.

#### S6.2 Explaining the shape of the curves in Fig. 4 of the main text

In this section, we provide more detailed explanations about the shapes of the establishment time curves depending on the mode of action of the drug. Throughout this discussion, it is

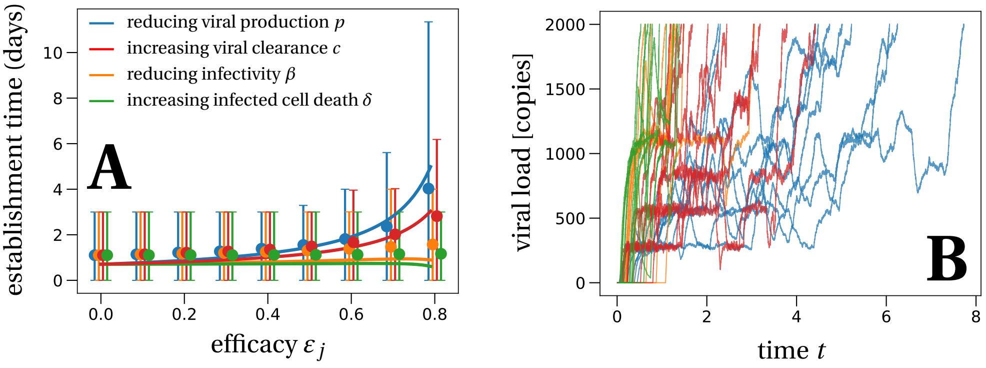

Figure S4: **The mean time to reach a detectable viral load at the infection site.** (This is Fig. 4 from the main text.) Panel A: Solid lines represent the theoretical prediction of the average time for the viral infection to reach 2,000 virions. We used the LowN parameter set to simulate 10,000 stochastic simulations that reached a viral load of 2,000 total virus particles when starting with an inoculum of $V_I(0) = 1$. Dots are the average times calculated from these simulations, error bars represent 90% of the simulated establishment times. Panel B: We plot 10 example trajectories that reach the detectable viral load for each of the four types of treatment (efficacy $\varepsilon_j = 0.75$). Under treatment that increases the infected cell death $\delta$, establishing trajectories reach the detectable viral load almost immediately. In contrast, drugs that directly affect the number of virus, i.e. clearance $c$ or production $p$, allow for trajectories that fluctuate much more, explaining the larger average detection times and the larger variation of detection times for these scenarios.

important to keep in mind that for the average establishment times, only trajectories that result in establishment are taken into account. To ease the discussion, Fig. S4 shows Fig. 4 from the main text.

Treatment that targets the virus infectivity $\beta$ does not increase the establishment time because these drugs do not affect the virus dynamics itself. Conditioned on virus establishment, the initially present virus particle will infect a target cell relatively quickly, i.e., on a similar time scale than without treatment, and then follow the same dynamics as without treatment. Since the burst size largely exceeds the detection threshold, in our model just two infected cells are sufficient to reach this threshold. Therefore, the establishment time remains largely unaffected by drugs targeting the infectivity $\beta$.

For drugs increasing the infected cell death rate $\delta$, the trajectories that contribute to the results in Fig. S4 are the ones that produced a large number of virus particles from a single cell in a short time. This is because of the strongly increased cell death rate for large values of efficacy $\varepsilon_\delta$. Therefore, a surviving virus trajectory needs to reach large numbers of virus particles in a short time to avoid extinction. This is different for a reduced viral production $p$ where the infected cell death rate is unaffected. Therefore, it is not necessary for a surviving virus trajectory to reach high viral loads very quickly, even though this is of course possible which is reflected by the large 90% confidence interval. This is visualized in Fig. S4, panel B: green trajectories correspond to drugs affecting the cell death rate and blue trajectories correspond to drugs reducing viral production.

Lastly, increasing the viral clearance rate $c$ by prophylactic treatment increases the establishment time with increasing efficacy, but not as much as treatment with drugs that reduce viral production $p$. The reason here is that clearance acts just after the viral production, i.e., there is time passing between the production of a virus particle and its clearance. Hence, reducing virus production has a stronger effect on the establishment time than an increase of viral clearance $c$ which acts later in the viral life cycle.

### S7 Parameter estimation

Patient data from Young et al. (2020) were fitted using the set of differential equations presented in eq. (1) in the main text. To ensure identifiability of critical parameters of the viral dynamics, i.e. the basic reproductive number $R_0$, the loss rate of infected cells $\delta$ and the viral production $p$, the remaining parameters $c$, $k$ and $V_0$ were fixed. Viral clearance $c$ was fixed to 10 day$^{-1}$. For the eclipse phase $k$ we chose 5 day$^{-1}$ and the initial inoculum $V_0$ was set to 1/30 copies.mL$^{-1}$ (see Gonçalves et al. (2020) for further details). Parameters were estimated in a non-linear mixed effect model using the SAEM algorithm implemented in Monolix (www.lixoft.com). The best fit using all available patient data resulted in the parameter values $R_0 = 7.69$, $\delta = 0.595$ and $p = 11,200$, the principal data set used in the main text (LowN parameter set).

#### S7.1 Parameter estimates for individuals plotted in Fig. 1 in the main text

Applying this method to data from four single patients in Young et al. (2020), we obtain the best parameter set for each individual. These individual parameter sets were used to plot the deterministic curves in Fig. 1 from the main text. The exact parameter values are given in Table S2.

| **Patient ID** | $R_0$ | $\delta$ | $p$ |
|---|---|---|---|
| 2 (blue) | 9.77 | 0.71 | 11,016 |
| 4 (orange) | 8.73 | 0.66 | 11,104 |
| 11 (red) | 9.2 | 0.86 | 11,060 |
| 18 (green) | 7.12 | 0.5 | 11,031 |

Table S2: **Model parameters used for the deterministic fits in Fig. 1 in the main text.** The other parameters are as stated in the Figure. The colors correspond to the line colors in the Figure.

#### S7.2 Sensitivity analysis with respect to variations in the fraction of infectious virus particles $\eta$

We evaluate how different choices of $\eta$, the fraction of infectious virus among all produced virus particles, affect the estimates of the within-host reproductive number $R_0$ and the burst size $\eta$. In the main text, we have used the parameter estimate with $\eta = 10^{-3}$ which resulted in $R_0 = 7.69$ and $N = 18,823$. For a larger fraction of infectious virus particles, $\eta = 10^{-2}$, we find $R_0 = 5.3$ and $N = 3,303$; for a smaller fraction of infectious virus particles, $\eta = 10^{-4}$, we obtain $R_0 = 9.2$ and $N = 349,367$. While the within-host reproductive number $R_0$ does not vary too much between the different choices of $\eta$, the burst size $N$ shows large variation. This has no effect on our results on the establishment of a SARS-CoV-2 infection because the burst size always enters in the form of a product with $\eta$. In all the different scenarios above, the product $\eta \times N$ varies between 18 for $\eta = 10^{-3}$ and 35 for $\eta = 10^{-4}$.

Overall, the differences in estimates for $R_0$ will affect the precise estimate of the critical efficacy and differences in the estimate for $N$ translate to differences in the quantitative values of the establishment probability curves below the critical efficacy. The predictions on the detection and extinction time strongly depend on the overall burst size $N$ so that these will vary considerably depending on the choice of $\eta$.

#### References

Anderson, D. F. & Kurtz, T. G. (2011), *Continuous Time Markov Chain Models for Chemical Reaction Networks*, pages 3–42. Springer New York, New York, NY. doi: 10.1007/978-1-4419-6766-4_1.

Bonhoeffer, S., May, R. M., Shaw, G. M., & Nowak, M. A. (1997). Virus dynamics and drug therapy. *Proceedings of the National Academy of Sciences*, 94(13):6971–6976. doi: 10.1073/pnas.94.13.6971.

Conway, J. M., Konrad, B. P., & Coombs, D. (2013). Stochastic analysis of pre- and postexposure prophylaxis against HIV infection. *SIAM Journal on Applied Mathematics*, 73(2):904–928. doi: 10.1137/120876800.

Desai, M. M. & Fisher, D. S. (2007). Beneficial mutation–selection balance and the effect of linkage on positive selection. *Genetics*, 176(3):1759–1798. doi: 10.1534/genetics.106.067678.

Duwal, S., Dickinson, L., Khoo, S., & von Kleist, M. (2019). Mechanistic framework predicts drug-class specific utility of antiretrovirals for HIV prophylaxis. *PLOS Computational Biology*, 15(1):e1006740. doi: 10.1371/journal.pcbi.1006740.

Gonçalves, A., Bertrand, J., Ke, R., Comets, E., de Lamballerie, X., Malvy, D., Pizzorno, A., Terrier, O., Calatrava, M. R., Mentré, F., Smith, P., Perelson, A. S., & Guedj, J. (2020). Timing of antiviral treatment initiation is critical to reduce sars-cov-2 viral load. *medRxiv*. doi: 10.1101/2020.04.04.20047886.

Haccou, P., Jagers, P., & Vatutin, V. A. (2005), *Branching Processes: Variation, Growth, and Extinction of Populations*. Cambridge Studies in Adaptive Dynamics. Cambridge University Press. doi: 10.1017/CBO9780511629136.

Hataye, J. M., Casazza, J. P., Best, K., Liang, C. J., Immonen, T. T., Ambrozak, D. R., Darko, S., Henry, A. R., Laboune, F., Maldarelli, F., Douek, D. C., Hengartner, N. W., Yamamoto, T., Keele, B. F., Perelson, A. S., & Koup, R. A. (2019). Principles governing establishment versus collapse of hiv-1 cellular spread. *Cell Host & Microbe*, 26(6):748 – 763.e20. doi: https://doi.org/10.1016/j.chom.2019.10.006.

Park, W. B., Kwon, N.-J., Choi, S.-J., Kang, C. K., Choe, P. G., Kim, J. Y., Yun, J., Lee, G.-W., Seong, M.-W., Kim, N. J., Seo, J.-S., & don Oh, M. (2020). Virus isolation from the first patient with SARS-CoV-2 in korea. *Journal of Korean Medical Science*, 35(7). doi: 10.3346/jkms.2020.35.e84.

Pearson, J. E., Krapivsky, P., & Perelson, A. S. (02 2011). Stochastic theory of early viral infection: Continuous versus burst production of virions. *PLOS Computational Biology*, 7(2):1–17. doi: 10.1371/journal.pcbi.1001058.

Young, B. E., Ong, S. W. X., Kalimuddin, S., Low, J. G., Tan, S. Y., Loh, J., Ng, O.-T., Marimuthu, K., Ang, L. W., Mak, T. M., Lau, S. K., Anderson, D. E., Chan, K. S., Tan, T. Y., Ng, T. Y., Cui, L.,

Said, Z., Kurupatham, L., Chen, M. I.-C., Chan, M., Vasoo, S., Wang, L.-F., Tan, B. H., Lin, R. T. P., Lee, V. J. M., Leo, Y.-S., & Lye, D. C. (2020). Epidemiologic Features and Clinical Course of Patients Infected With SARS-CoV-2 in Singapore. *JAMA*. doi: 10.1001/jama.2020.3204.
